# Safety Monitoring of Health Outcomes following Influenza Vaccination during the 2023-2024 Season among U.S. Medicare Beneficiaries Aged 65 Years and Older

**DOI:** 10.1101/2025.01.03.25319959

**Authors:** Patricia C. Lloyd, Gyanada Acharya, Henu Zhao, Nimesh Shah, Godwin Anguzu, Derick Ambarsoomzadeh, Tainya C. Clarke, Xinyi Ng, Mao Hu, Yoganand Chillarige, Richard A. Forshee, Steven A. Anderson

**Author notes:** **Corresponding Author: Patricia C. Lloyd, PhD, ScM** Health Statistician Office of Biostatistics and Pharmacovigilance Center for Biologics Evaluation and Research U.S. Food & Drug Administration 10903 New Hampshire Ave., Building 71 Silver Spring, MD 20993. **Author Email Addresses:**.

## Abstract

Background

Influenza vaccination is widely recommended for individuals aged 6 months and older in the United States (U.S.). While the safety of annual influenza vaccines is well established, FDA conducts routine monitoring and evaluation of safety. This study assessed the safety of 2023-2024 influenza vaccines among elderly U.S. Medicare beneficiaries.

Methods

A self-controlled case series (SCCS) analysis compared incidence rate ratios (IRR) of anaphylaxis, encephalitis/encephalomyelitis/acute disseminated encephalomyelitis, Guillain-Barré syndrome (GBS), transverse myelitis, hemorrhagic stroke (HS), non-hemorrhagic stroke (NHS), transient ischemic attack (TIA), and NHS or TIA, following 2023-2024 seasonal influenza vaccinations in risk and control intervals among Medicare beneficiaries aged 65 years and older. We used conditional Poisson regression to estimate IRRs and 95% confidence intervals (CIs) adjusted for event-dependent observation time for certain outcomes, seasonality, and uncertainty from outcome misclassification where feasible. For health outcomes with statistically significant associations, we stratified results by concomitant vaccination status.

Results

We observed a total of 20,258,006 influenza vaccinees among the Medicare population, and no statistically significant elevations of risk for anaphylaxis, encephalitis/encephalomyelitis (with ADEM), GBS, HS, or TM. For the combined NHS/TIA outcome (22-42-day risk window), we observed a small elevation in risk that was statistically significant in both the Fee-For-Service (FFS) and Medicare Advantage (MA) populations that received a high-dose vaccine. This risk was also statistically significant among MA beneficiaries that received any influenza vaccine. Additionally, we observed a small statistically significant risk for the individual TIA outcome (22-42-day risk window) among the MA population that received any influenza vaccine.

Conclusion

Results from this study indicate that the benefits of seasonal influenza vaccines continue to outweigh the risks. The small statistically significant increased risk of stroke outcomes observed in the study must be carefully considered in light of the known benefits of influenza vaccination.

## INTRODUCTION

The Centers for Disease Control and Prevention’s (CDC) Advisory Committee on Immunization Practices (ACIP) strongly recommends influenza vaccination annually for all individuals aged 6 months and older in the United States (U.S.), recognizing its role as a cornerstone for primary prevention against seasonal influenza.^(1)^ Influenza vaccines are reformulated each year to target the expected dominant circulating strains, and the U.S. Food and Drug Administration’s (FDA) Vaccine and Related Biological Products Advisory Committee (VRBPAC) recommended the 2023-2024 season’s vaccine composition to remain relatively consistent with the prior season.^(2,3)^

ACIP recommended three influenza vaccines for use in people aged 65 years or older: High-dose inactivated influenza vaccines (HD-IIV4s), recombinant influenza vaccines (RIV4s), and the quadrivalent adjuvanted inactivated influenza vaccine (aIIV4).^(2)^ For the 2023-2024 season, multiple IIV4s and RIV4s were available, each with age-specific guidelines (Supplemental Table 1). The Afluria, Fluarix, FluLaval, Fluzone, and Flucelvax Quadrivalent vaccines were approved for people ages 6 months and older, and the Flublok Quadrivalent vaccine was approved for people ages 18 years and older.^(4, 5)^

While influenza vaccines are generally considered safe based on evidence from past seasons, surveillance is useful for monitoring for potential associations with specific adverse health outcomes such as anaphylaxis, encephalitis/encephalomyelitis, Guillain-Barré syndrome (GBS), stroke, and transverse myelitis.^(6, 7)^ Safety monitoring can identify and address any emerging concerns associated with alterations in vaccine formulations, particularly considering the widespread use of influenza vaccines in the Medicare population. This includes populations with weak immune responses to the influenza vaccine and those with pre-existing conditions, such as obesity or diabetes.^(8)^

The U.S. FDA Center for Biologics Evaluation and Research (CBER) monitors the safety of vaccines and conducted surveillance of the 2023-2024 season’s influenza vaccines for several potential health outcomes. Through a collaborative agreement between FDA and the Centers for Medicare & Medicaid Services (CMS), we conducted a self-controlled case series (SCCS) study to assess the safety of the 2023–2024 seasonal influenza vaccines among Medicare beneficiaries aged 65 years and older. The primary objectives of this study were to: i) monitor influenza vaccination uptake for the 2023–2024 season and calculate observed counts and rates for outcomes of interest and ii) use the SCCS method to estimate the incidence rate ratio (IRR) and examine the association between influenza vaccination and each pre-specified health outcome, when sufficiently powered. The secondary objective of this study was to examine the effect of concomitant vaccination, when case counts allowed, on the risk of health outcomes with statistically significant associations in the primary analyses.

## METHODS

### 2.1 Data Sources, Study Population, and Study Period

This study utilized administrative claims data from CMS. The CMS Medicare claims database offers comprehensive data on healthcare service usage, sourced from Medicare Shared Systems Data (SSD) for Fee-for-Service (FFS; Medicare Parts A and B, but not Part C) enrollees and the Encounter Data System (EDS) for Medicare Advantage (MA) enrollees, covering inpatient, outpatient, and pharmacy settings (Medicare Part D claims). We used the CMS Medicare Enrollment Database (EDB) and Common Medicare Environment (CME) to capture demographic characteristics and death information, and the Minimum Data Set (MDS) 3.0 to determine nursing home residency status.

This study included individuals aged 65 years and older in the FFS and MA Medicare populations who received a 2023–2024 influenza vaccine and met eligibility criteria outlined in Supplementary Figure 1. The study start date was August 1, 2023, which aligned with the start of the 2023–2024 influenza season in the US. We had two separate study end dates for the two Medicare populations, June 29, 2024 (FFS) and August 3, 2024 (MA), to ensure adequate vaccine capture and 90% data completeness for health outcomes of interest.^(9)^

### 2.2 Exposure and Outcomes

The exposure in this study was the first influenza vaccine administration observed during the study period. Exposures for influenza vaccines (Supplemental Table 2) were identified using vaccine-specific codes, including Current Procedural Terminology (CPT), Healthcare Common Procedure Coding System (HCPCS), and National Drug Codes (NDCs) in the inpatient, outpatient professional, professional, or pharmacy care settings. Influenza vaccines included the HD-IIV4 and the quadrivalent aIIV4. All other vaccine types, including indication of general influenza vaccine administration, were grouped into one category.

Our study monitored eight incident outcomes: anaphylaxis, encephalitis/encephalomyelitis/acute disseminated encephalomyelitis, GBS, transverse myelitis (TM), hemorrhagic stroke (HS), non-hemorrhagic stroke (NHS), transient ischemic attack (TIA), and a composite NHS or TIA outcome (NHS/TIA). These were captured in the observation period following influenza vaccination, and identified using *International Classification of Diseases, Tenth Revision, Clinical Modification* codes in either primary or secondary diagnosis positions (except GBS, which was assessed in only the primary diagnosis position) (Supplemental Table 3). To be classified as an incident outcome, a particular outcome must not have prior occurrences during a specified time period (i.e., the clean window). Additional outcome-specific exclusions were applied for HS, NHS, NHS/TIA, TIA, and TM (Supplemental Table 4).

### 2.3 Covariates

To characterize the study population, we defined various demographic, socioeconomic, geographic, and medical covariates. We used Medicare enrollment databases to obtain information on demographic, socioeconomic and geographic characteristics, and diagnosis or procedure codes from medical claims to identify concomitant vaccinations, immunocompromising and medical conditions. We assessed medical conditions in the 365 days prior to the influenza vaccination, and concomitant vaccinations on the same day as the influenza vaccination.

### 2.4 Study Design and Follow-up

We used an SCCS design to compare incident health outcomes during post-vaccination risk and control intervals, controlling for time-invariant confounders by making comparisons within, rather than between, individuals.^(10)^ Individuals were followed for the duration of the outcome-specific observation period, starting from the date of influenza vaccination and ending 90 days thereafter for all outcomes except anaphylaxis (16 days post-vaccination). The risk interval was a period of hypothesized excess risk following vaccination and was defined for each outcome based on literature review and clinician input and has been used in several prior FDA studies.^(11, 12)^ For our sensitivity analyses, a washout period, defined as the period between the risk and control interval, was established to avoid carryover effects from vaccination contributing to the control interval. The control interval comprised all other time in the post-vaccination observation period not in the risk interval or washout period. The claims settings, risk and control intervals, and clean windows for each outcome are defined in Supplemental Table 3. Individuals’ time intervals were censored at disenrollment, death, receipt of a subsequent influenza vaccine, end of the observation period, or the end of the study period.

### 2.5 Statistical Analysis

#### 2.5.1 Descriptive Analyses

We produced descriptive summaries of vaccine recipient characteristics, vaccine uptake, and health outcomes following influenza vaccination. Additionally, we generated a demographic and medical covariate-stratified outcome summary describing vaccination counts, outcome counts, and outcome rates (per 100,000 person-years) within individual outcome cohorts.

#### 2.5.2 Inferential Analyses

The primary SCCS analysis was conducted for individuals who received any age-appropriate influenza vaccine and estimated IRRs and 95% CIs, comparing health outcome rates in the risk and control intervals using a conditional Poisson regression. For each health outcome, Attributable Risk (AR) was calculated per 100,000 vaccinations and person-years, with a standard error estimated through bootstrap resampling. Secondary analysis was conducted only for health outcomes with a statistically significant increased IRR observed in the primary analysis and consisted of two subgroups: (1) those with an observed influenza vaccination on the same day as any prespecified concomitant vaccine, and (2) those with an observed influenza vaccination without any prespecified concomitant vaccine.^a^

The primary and secondary analyses were adjusted for (i) event-dependent observation time for outcomes with 30-day case fatality rates (CFR) of 10 percent or higher, (ii) seasonality for all outcomes, and (iii) positive predictive value (PPV) for outcomes with available PPV from prior medical record reviews. The adjustment for event-dependent observation time, developed by Farrington et al. (herein referred to as the “Farrington adjustment”), was applied to outcomes with a CFR of 10 percent or higher, compensating for shortened observation times.^(13)^ The CFR was calculated in the FFS SCCS study population by measuring the percentage of deaths within 30 days of an incident outcome in 2022.^b^ To reduce potential bias from seasonal outcome patterns, we adjusted for seasonality by using incidence rates of outcomes estimated from the full Medicare population 65 years and older from corresponding calendar months in 2022-2023 as the baseline incidence of outcomes in the risk and control intervals. To address potential misclassification from claims-based outcome definitions, quantitative bias analysis with multiple imputation was performed for outcomes with available PPV estimates from prior studies (Supplemental Table 5). The PPV indicates the proportion of true cases among those verified through medical record reviews from prior vaccine safety studies. Using PPV-based multiple imputation, we addressed uncertainty from outcome misclassification by generating simulated datasets where cases were sampled with probabilities equal to the PPVs derived from prior medical record review in the FFS population. PPV-based imputation analyses could not be applied across all health outcomes because certain outcomes were not able to be included in medical record reviews and did not have available estimates from previous studies. Supplemental Table 6 presents the most adjusted primary analysis for each outcome.

Sensitivity analyses were performed for the primary analysis. First, a washout period of 14-days between risk and control intervals (applicable for all outcomes except anaphylaxis) was added to reduce potential bias from carryover effects from vaccination contributing to control interval risk. Second, for outcomes with a case fatality rate ≥ 10%, the full planned observation period was credited to each individual, regardless of death or disenrollment. The Farrington adjustment was removed to evaluate potential bias stemming from violation of the SCCS assumption regarding the independence of events from observation length.^(13)^

All analyses were conducted using R 4.3.2 (R Foundation for Statistical Computing, Vienna, Austria), and SAS v.9.4 (SAS Institute Inc., Cary, NC, United States). This surveillance activity was conducted as part of the FDA public health surveillance mandate, and the surveillance activities are not considered research and therefore are exempt from institutional review board review and approval.^(14)^

## RESULTS

### 3.1 Descriptive Results

We observed a total of 20,258,006 influenza vaccinees among the Medicare population, with 12,660,709 FFS and 7,597,297 MA beneficiaries receiving a vaccine during the study period (Figures 1a and 1b). The HD-IIV4 (high-dose) vaccine accounted for about half of the total uptake in both the FFS (49.5%) and MA (54.4%) populations, while the aIIV4 (adjuvanted) vaccine was received by 41.9% and 26.8% of FFS and MA beneficiaries, respectively. Approximately 8.6% of FFS and 18.9% of MA beneficiaries received other vaccine type. Vaccine distribution was relatively consistent across various socio-demographic and geographical characteristics in both the FFS and MA populations (Table 1). The majority of Medicare FFS (72%) and MA (73%) vaccinated individuals were aged 65-79 years. And, in both the vaccinated FFS and MA populations, there was a higher proportion of females (∼58.0%) compared to males (∼42.0%). The majority of vaccinees were white (86.7% in FFS and 72.9% in MA), and majority of our study population were community-dwelling (i.e., did not reside in a nursing home) (98.0% in FFS and 98.3% in MA). We also observed concomitant vaccinations among the study population, with COVID-19 (21.8% in FFS and 0.6% in MA) and the respiratory syncytial virus (RSV) (7.0% in FFS and 0.9% in MA) being the most common vaccines administered with influenza.

**Figure 1a.**
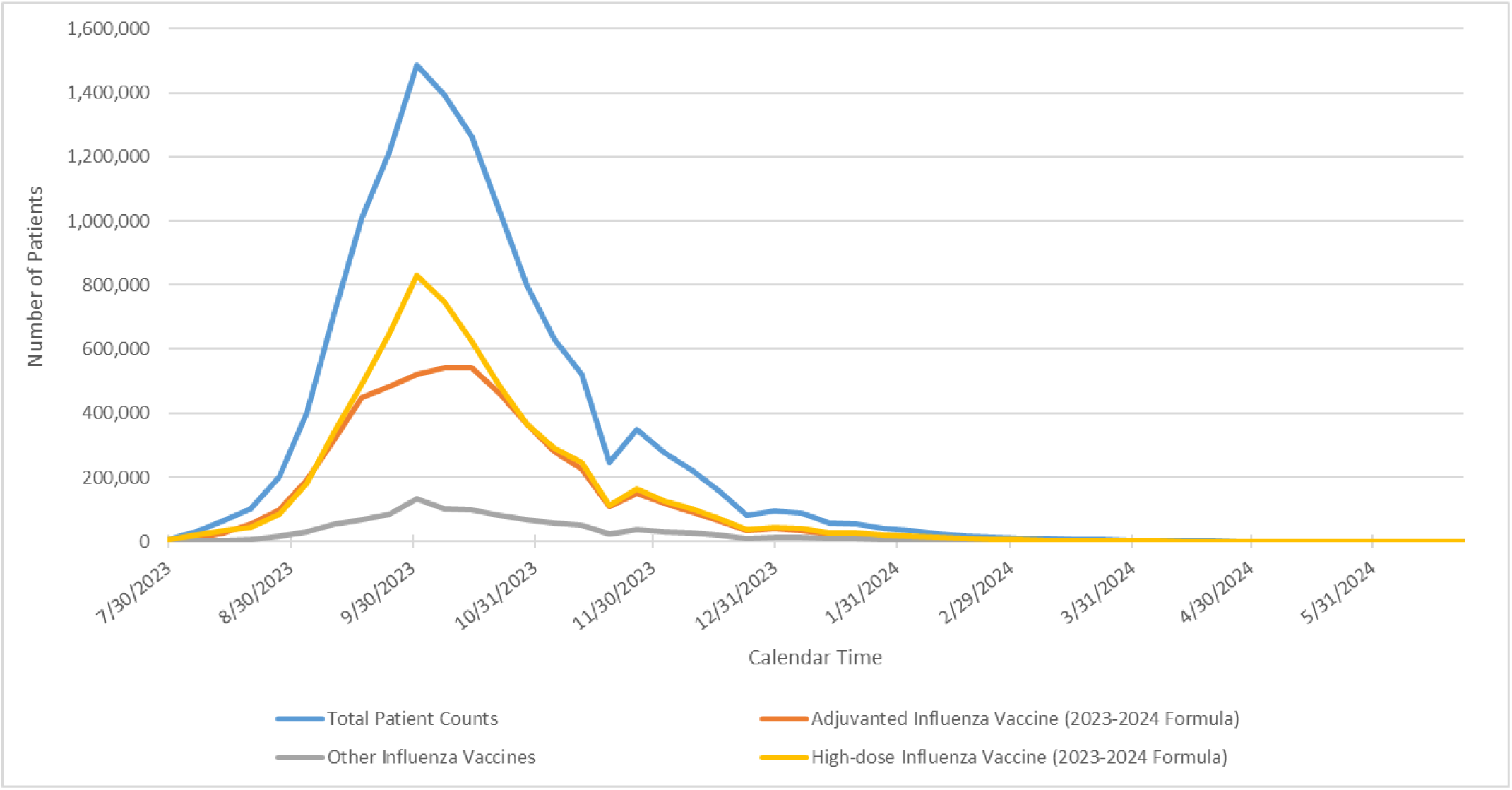
Number of Vaccinated Patients with Qualifying Influenza Vaccine by Month; Total and by Vaccine Type, Fee-For-Service (data through 6/29/2024)

**Figure 1b.**
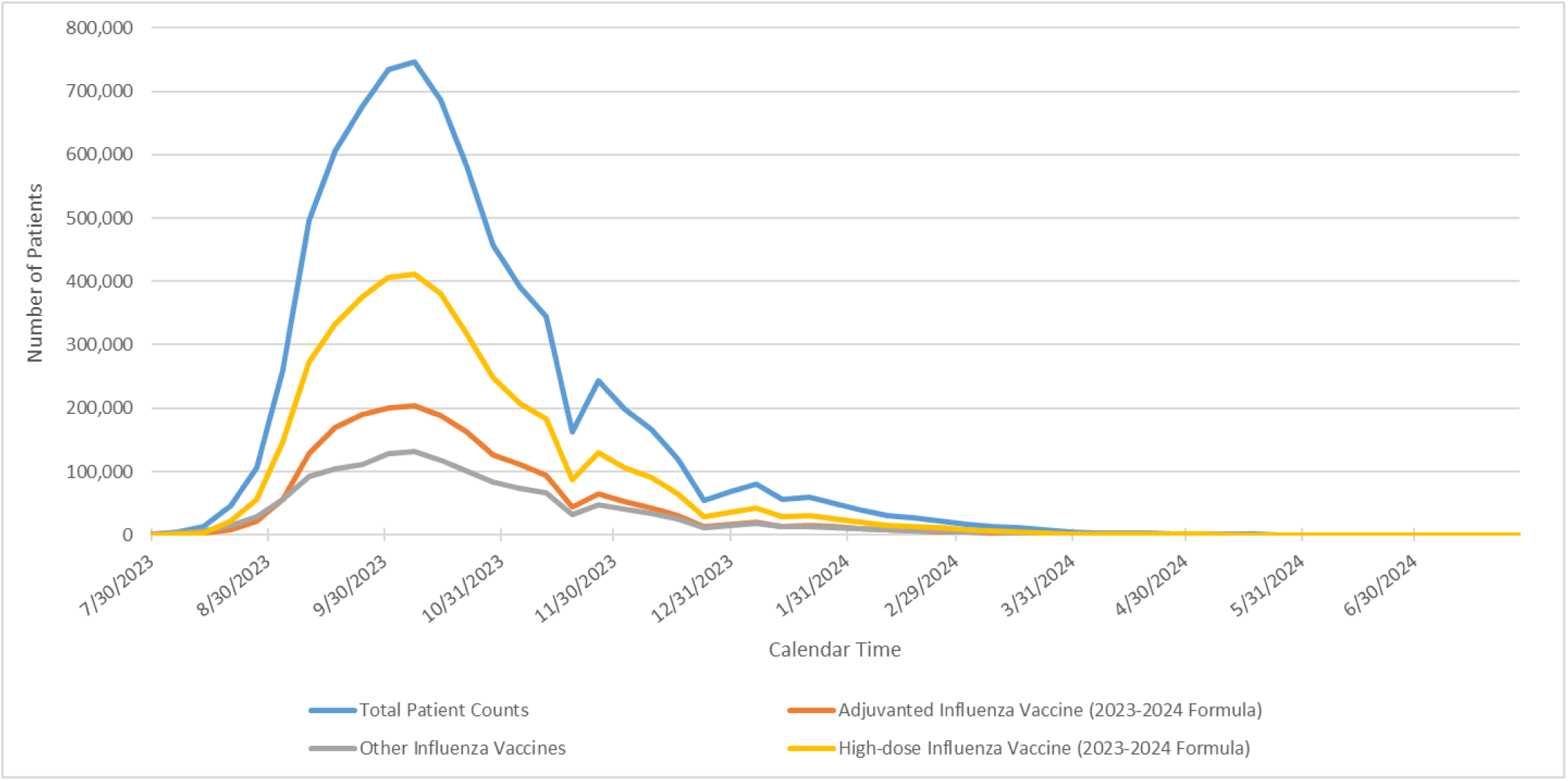
Number of Vaccinated Patients with Qualifying Influenza Vaccine by Month; Total and by Vaccine Type, Medicare Advantage (data through 8/2/2024)

**Figure 2a.**
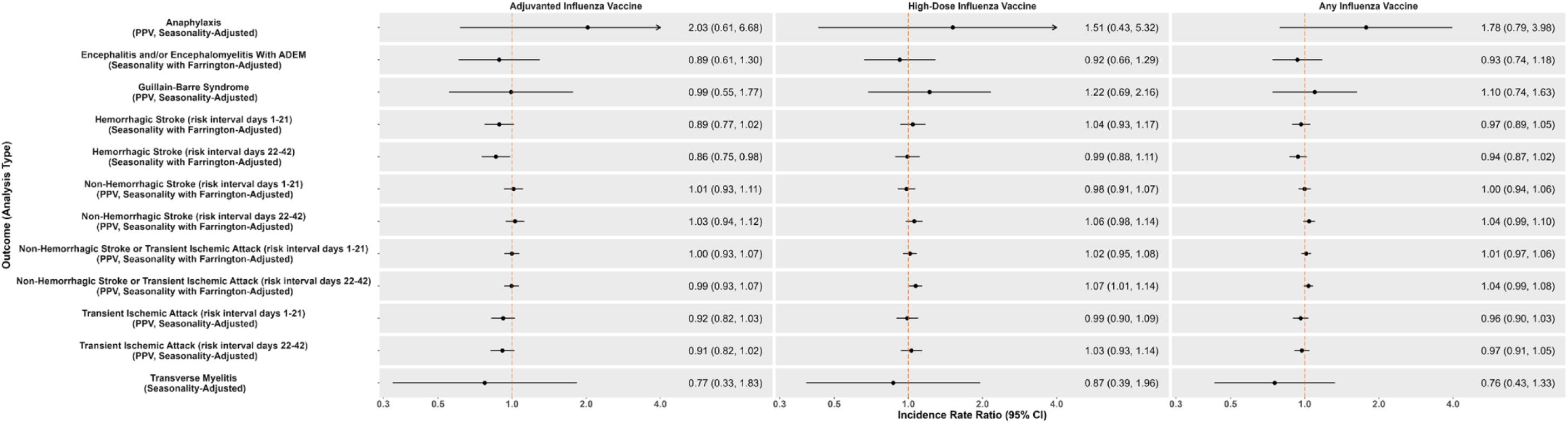
Forest Plot of Incidence Rate Ratios (95% CI) in Primary SCCS by Analysis and Vaccine Type, Fee-For-Service

**Figure 2b.**
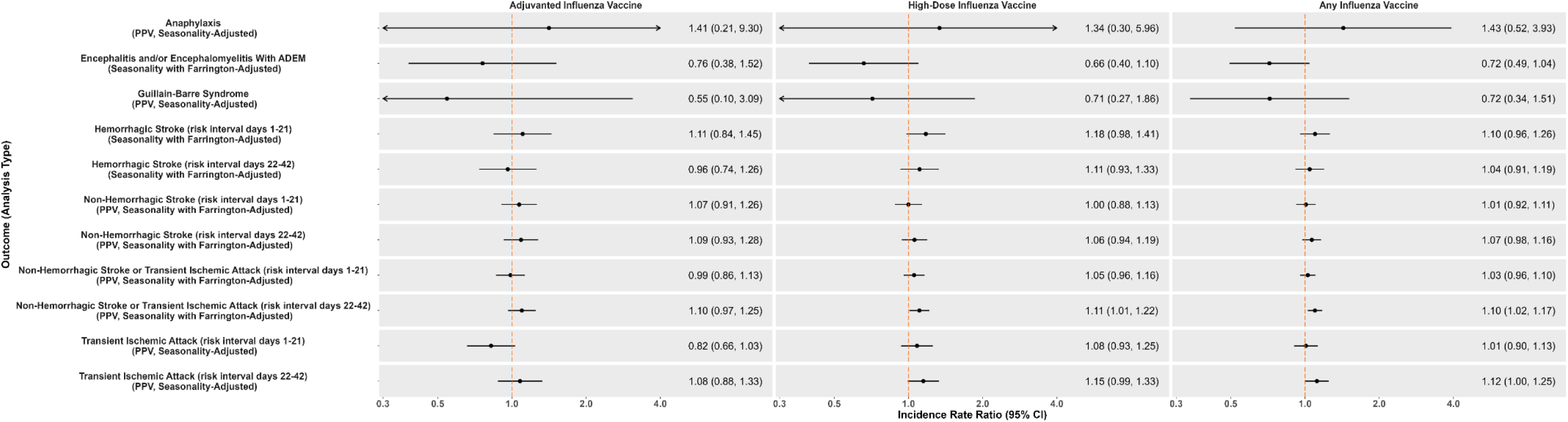
Forest Plot of Incidence Rate Ratios (95% CI) in Primary SCCS by Analysis and Vaccine Type, Medicare Advantage

**Table 1.**
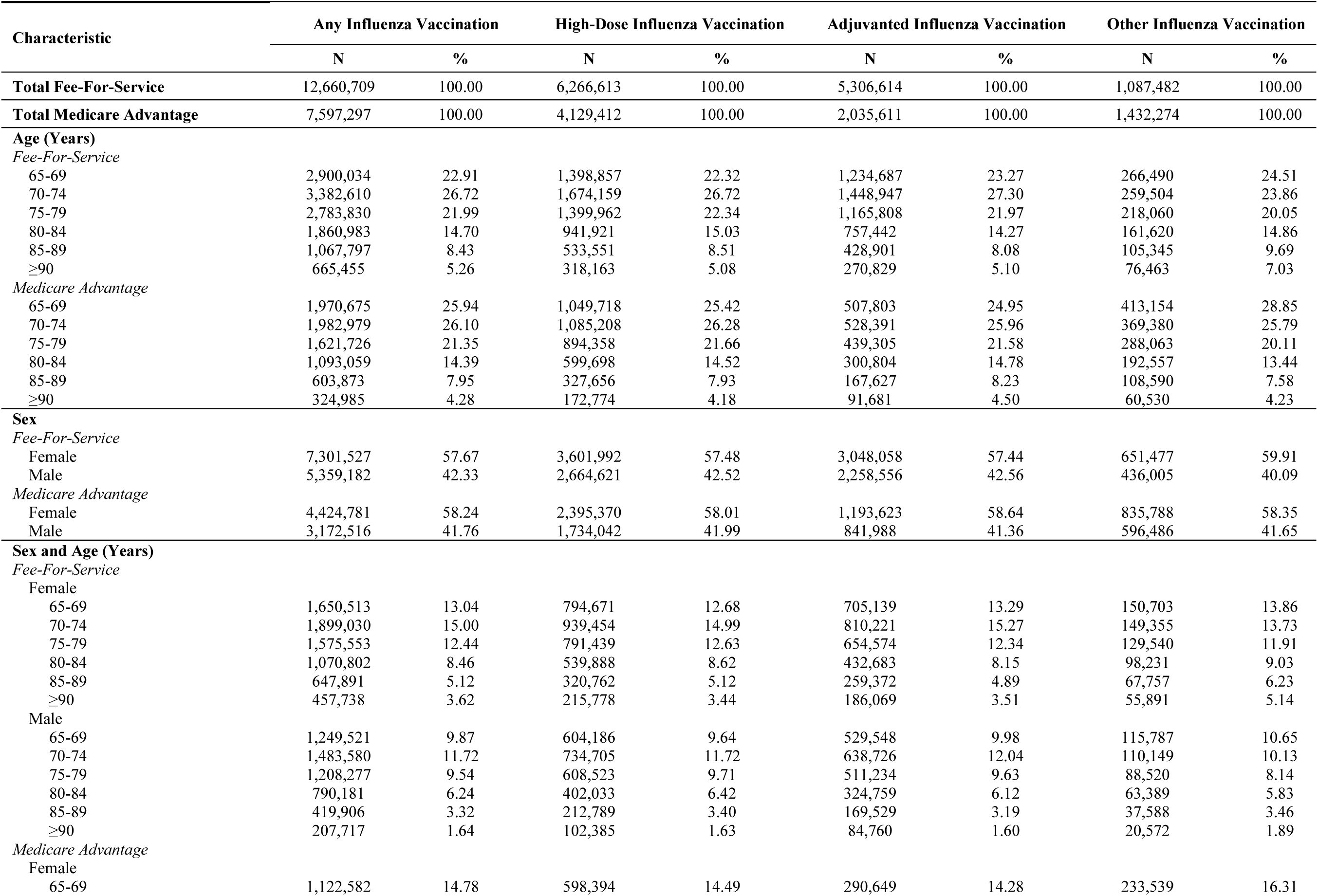
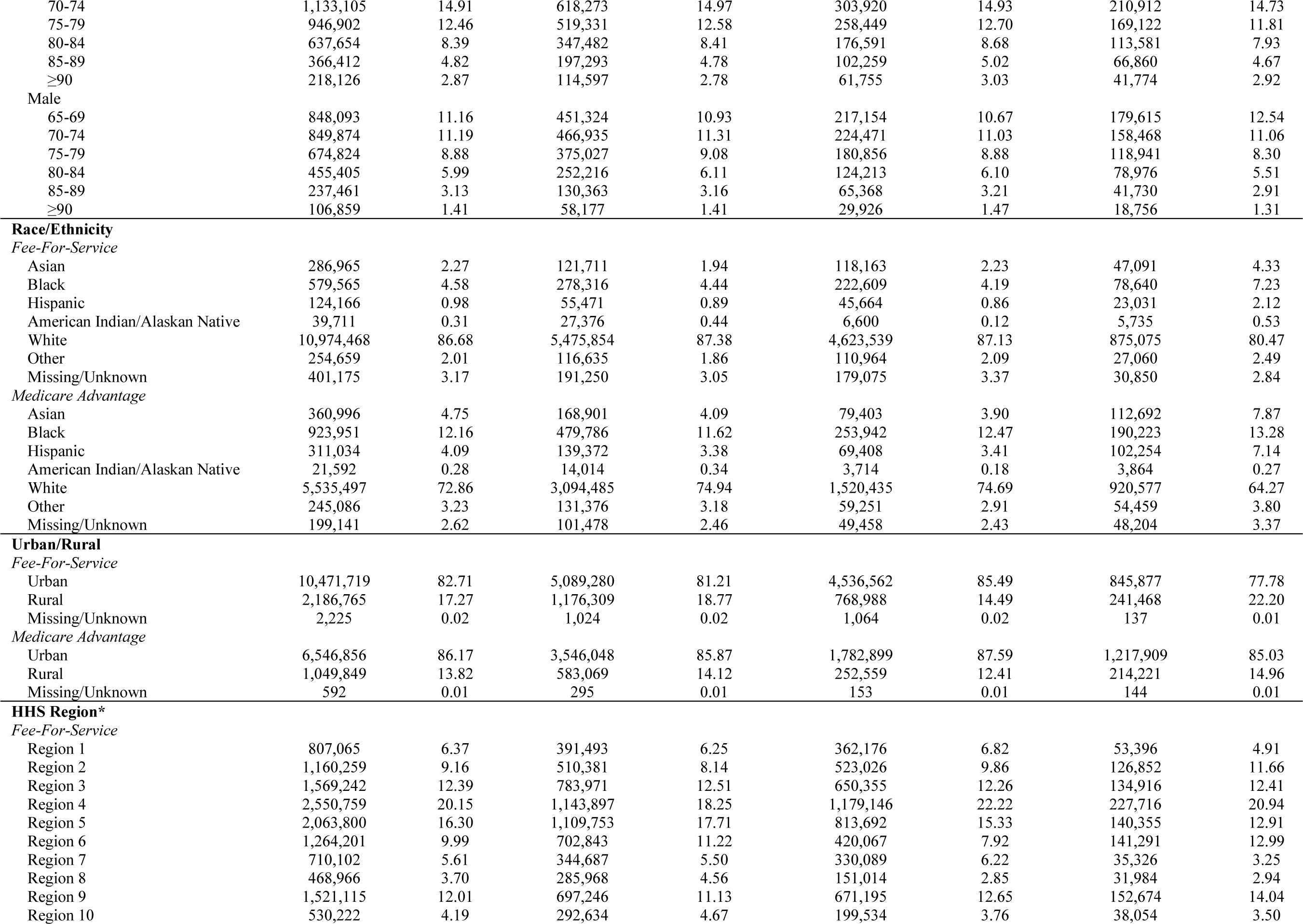
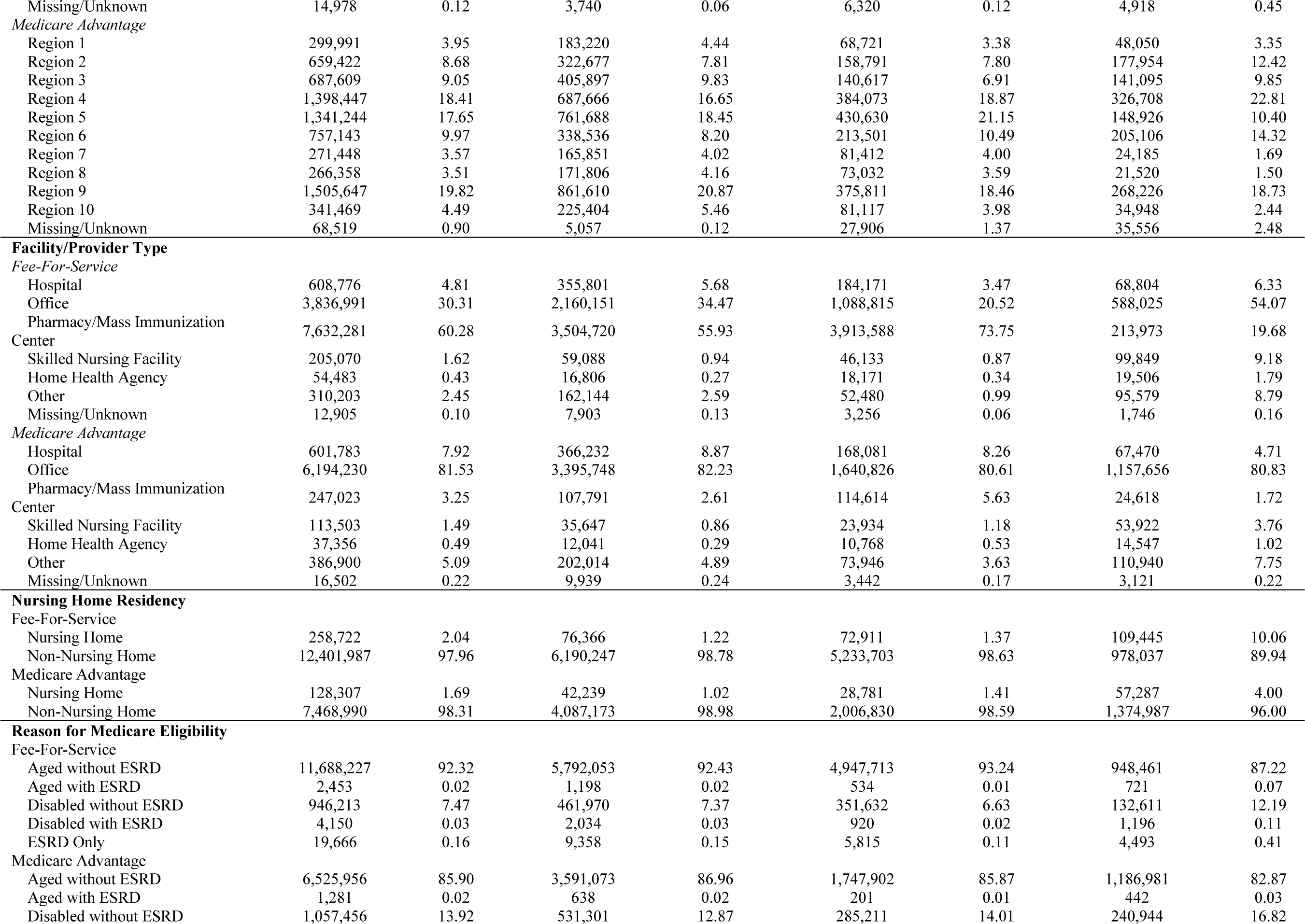
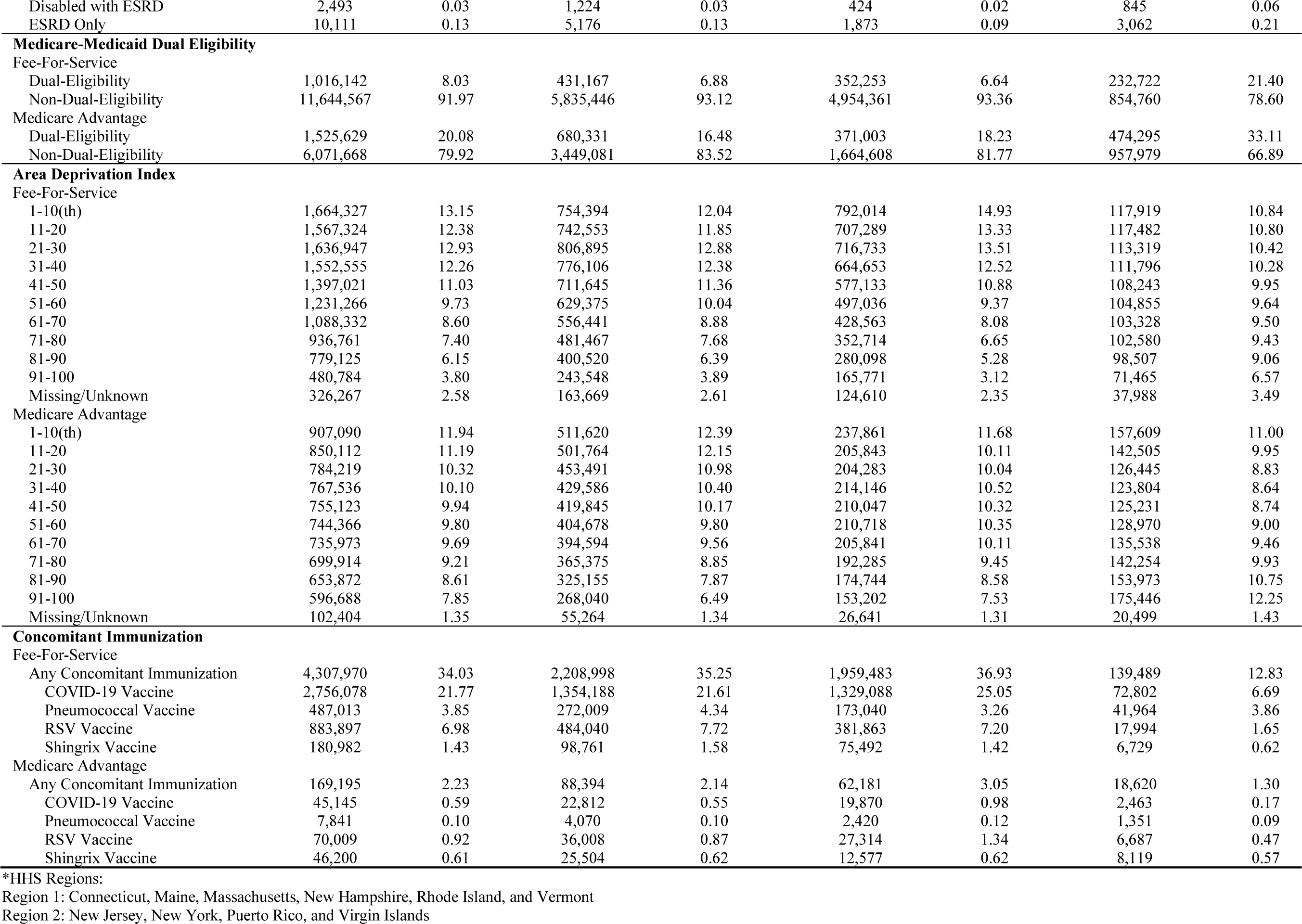
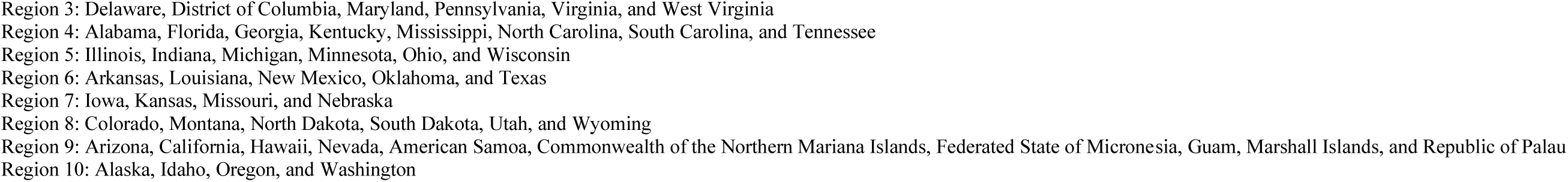
Socio-demographic and Geographical Characteristics of Persons Receiving Influenza Vaccines Stratified by Vaccine Type and Medicare Plan.

| Characteristic | Any Influenza Vaccination |  | High-Dose Influenza Vaccination |  | Adjuvanted Influenza Vaccination |  | Other Influenza Vaccination |  |
| --- | --- | --- | --- | --- | --- | --- | --- | --- |
|  | N | % | N | % | N | % | N | % |
| **Total Fee-For-Service** | 12,660,709 | 100.00 | 6,266,613 | 100.00 | 5,306,614 | 100.00 | 1,087,482 | 100.00 |
| **Total Medicare Advantage** | 7,597,297 | 100.00 | 4,129,412 | 100.00 | 2,035,611 | 100.00 | 1,432,274 | 100.00 |
| **Age (Years)** |  |  |  |  |  |  |  |  |
| *Fee-For-Service* |  |  |  |  |  |  |  |  |
| 65-69 | 2,900,034 | 22.91 | 1,398,857 | 22.32 | 1,234,687 | 23.27 | 266,490 | 24.51 |
| 70-74 | 3,382,610 | 26.72 | 1,674,159 | 26.72 | 1,448,947 | 27.30 | 259,504 | 23.86 |
| 75-79 | 2,783,830 | 21.99 | 1,399,962 | 22.34 | 1,165,808 | 21.97 | 218,060 | 20.05 |
| 80-84 | 1,860,983 | 14.70 | 941,921 | 15.03 | 757,442 | 14.27 | 161,620 | 14.86 |
| 85-89 | 1,067,797 | 8.43 | 533,551 | 8.51 | 428,901 | 8.08 | 105,345 | 9.69 |
| ≥90 | 665,455 | 5.26 | 318,163 | 5.08 | 270,829 | 5.10 | 76,463 | 7.03 |
| *Medicare Advantage* |  |  |  |  |  |  |  |  |
| 65-69 | 1,970,675 | 25.94 | 1,049,718 | 25.42 | 507,803 | 24.95 | 413,154 | 28.85 |
| 70-74 | 1,982,979 | 26.10 | 1,085,208 | 26.28 | 528,391 | 25.96 | 369,380 | 25.79 |
| 75-79 | 1,621,726 | 21.35 | 894,358 | 21.66 | 439,305 | 21.58 | 288,063 | 20.11 |
| 80-84 | 1,093,059 | 14.39 | 599,698 | 14.52 | 300,804 | 14.78 | 192,557 | 13.44 |
| 85-89 | 603,873 | 7.95 | 327,656 | 7.93 | 167,627 | 8.23 | 108,590 | 7.58 |
| ≥90 | 324,985 | 4.28 | 172,774 | 4.18 | 91,681 | 4.50 | 60,530 | 4.23 |
| **Sex** |  |  |  |  |  |  |  |  |
| *Fee-For-Service* |  |  |  |  |  |  |  |  |
| Female | 7,301,527 | 57.67 | 3,601,992 | 57.48 | 3,048,058 | 57.44 | 651,477 | 59.91 |
| Male | 5,359,182 | 42.33 | 2,664,621 | 42.52 | 2,258,556 | 42.56 | 436,005 | 40.09 |
| *Medicare Advantage* |  |  |  |  |  |  |  |  |
| Female | 4,424,781 | 58.24 | 2,395,370 | 58.01 | 1,193,623 | 58.64 | 835,788 | 58.35 |
| Male | 3,172,516 | 41.76 | 1,734,042 | 41.99 | 841,988 | 41.36 | 596,486 | 41.65 |
| **Sex and Age (Years)** |  |  |  |  |  |  |  |  |
| *Fee-For-Service* |  |  |  |  |  |  |  |  |
| Female |  |  |  |  |  |  |  |  |
| 65-69 | 1,650,513 | 13.04 | 794,671 | 12.68 | 705,139 | 13.29 | 150,703 | 13.86 |
| 70-74 | 1,899,030 | 15.00 | 939,454 | 14.99 | 810,221 | 15.27 | 149,355 | 13.73 |
| 75-79 | 1,575,553 | 12.44 | 791,439 | 12.63 | 654,574 | 12.34 | 129,540 | 11.91 |
| 80-84 | 1,070,802 | 8.46 | 539,888 | 8.62 | 432,683 | 8.15 | 98,231 | 9.03 |
| 85-89 | 647,891 | 5.12 | 320,762 | 5.12 | 259,372 | 4.89 | 67,757 | 6.23 |
| ≥90 | 457,738 | 3.62 | 215,778 | 3.44 | 186,069 | 3.51 | 55,891 | 5.14 |
| Male |  |  |  |  |  |  |  |  |
| 65-69 | 1,249,521 | 9.87 | 604,186 | 9.64 | 529,548 | 9.98 | 115,787 | 10.65 |
| 70-74 | 1,483,580 | 11.72 | 734,705 | 11.72 | 638,726 | 12.04 | 110,149 | 10.13 |
| 75-79 | 1,208,277 | 9.54 | 608,523 | 9.71 | 511,234 | 9.63 | 88,520 | 8.14 |
| 80-84 | 790,181 | 6.24 | 402,033 | 6.42 | 324,759 | 6.12 | 63,389 | 5.83 |
| 85-89 | 419,906 | 3.32 | 212,789 | 3.40 | 169,529 | 3.19 | 37,588 | 3.46 |
| ≥90 | 207,717 | 1.64 | 102,385 | 1.63 | 84,760 | 1.60 | 20,572 | 1.89 |
| *Medicare Advantage* |  |  |  |  |  |  |  |  |
| Female |  |  |  |  |  |  |  |  |
| 65-69 | 1,122,582 | 14.78 | 598,394 | 14.49 | 290,649 | 14.28 | 233,539 | 16.31 |
| 70-74 | 1,133,105 | 14.91 | 618,273 | 14.97 | 303,920 | 14.93 | 210,912 | 14.73 |
| 75-79 | 946,902 | 12.46 | 519,331 | 12.58 | 258,449 | 12.70 | 169,122 | 11.81 |
| 80-84 | 637,654 | 8.39 | 347,482 | 8.41 | 176,591 | 8.68 | 113,581 | 7.93 |
| 85-89 | 366,412 | 4.82 | 197,293 | 4.78 | 102,259 | 5.02 | 66,860 | 4.67 |
| ≥90 | 218,126 | 2.87 | 114,597 | 2.78 | 61,755 | 3.03 | 41,774 | 2.92 |
| Male |  |  |  |  |  |  |  |  |
| 65-69 | 848,093 | 11.16 | 451,324 | 10.93 | 217,154 | 10.67 | 179,615 | 12.54 |
| 70-74 | 849,874 | 11.19 | 466,935 | 11.31 | 224,471 | 11.03 | 158,468 | 11.06 |
| 75-79 | 674,824 | 8.88 | 375,027 | 9.08 | 180,856 | 8.88 | 118,941 | 8.30 |
| 80-84 | 455,405 | 5.99 | 252,216 | 6.11 | 124,213 | 6.10 | 78,976 | 5.51 |
| 85-89 | 237,461 | 3.13 | 130,363 | 3.16 | 65,368 | 3.21 | 41,730 | 2.91 |
| ≥90 | 106,859 | 1.41 | 58,177 | 1.41 | 29,926 | 1.47 | 18,756 | 1.31 |
| **Race/Ethnicity** |  |  |  |  |  |  |  |  |
| *Fee-For-Service* |  |  |  |  |  |  |  |  |
| Asian | 286,965 | 2.27 | 121,711 | 1.94 | 118,163 | 2.23 | 47,091 | 4.33 |
| Black | 579,565 | 4.58 | 278,316 | 4.44 | 222,609 | 4.19 | 78,640 | 7.23 |
| Hispanic | 124,166 | 0.98 | 55,471 | 0.89 | 45,664 | 0.86 | 23,031 | 2.12 |
| American Indian/Alaskan Native | 39,711 | 0.31 | 27,376 | 0.44 | 6,600 | 0.12 | 5,735 | 0.53 |
| White | 10,974,468 | 86.68 | 5,475,854 | 87.38 | 4,623,539 | 87.13 | 875,075 | 80.47 |
| Other | 254,659 | 2.01 | 116,635 | 1.86 | 110,964 | 2.09 | 27,060 | 2.49 |
| Missing/Unknown | 401,175 | 3.17 | 191,250 | 3.05 | 179,075 | 3.37 | 30,850 | 2.84 |
| *Medicare Advantage* |  |  |  |  |  |  |  |  |
| Asian | 360,996 | 4.75 | 168,901 | 4.09 | 79,403 | 3.90 | 112,692 | 7.87 |
| Black | 923,951 | 12.16 | 479,786 | 11.62 | 253,942 | 12.47 | 190,223 | 13.28 |
| Hispanic | 311,034 | 4.09 | 139,372 | 3.38 | 69,408 | 3.41 | 102,254 | 7.14 |
| American Indian/Alaskan Native | 21,592 | 0.28 | 14,014 | 0.34 | 3,714 | 0.18 | 3,864 | 0.27 |
| White | 5,535,497 | 72.86 | 3,094,485 | 74.94 | 1,520,435 | 74.69 | 920,577 | 64.27 |
| Other | 245,086 | 3.23 | 131,376 | 3.18 | 59,251 | 2.91 | 54,459 | 3.80 |
| Missing/Unknown | 199,141 | 2.62 | 101,478 | 2.46 | 49,458 | 2.43 | 48,204 | 3.37 |
| **Urban/Rural** |  |  |  |  |  |  |  |  |
| *Fee-For-Service* |  |  |  |  |  |  |  |  |
| Urban | 10,471,719 | 82.71 | 5,089,280 | 81.21 | 4,536,562 | 85.49 | 845,877 | 77.78 |
| Rural | 2,186,765 | 17.27 | 1,176,309 | 18.77 | 768,988 | 14.49 | 241,468 | 22.20 |
| Missing/Unknown | 2,225 | 0.02 | 1,024 | 0.02 | 1,064 | 0.02 | 137 | 0.01 |
| *Medicare Advantage* |  |  |  |  |  |  |  |  |
| Urban | 6,546,856 | 86.17 | 3,546,048 | 85.87 | 1,782,899 | 87.59 | 1,217,909 | 85.03 |
| Rural | 1,049,849 | 13.82 | 583,069 | 14.12 | 252,559 | 12.41 | 214,221 | 14.96 |
| Missing/Unknown | 592 | 0.01 | 295 | 0.01 | 153 | 0.01 | 144 | 0.01 |
| **HHS Region\*** |  |  |  |  |  |  |  |  |
| *Fee-For-Service* |  |  |  |  |  |  |  |  |
| Region 1 | 807,065 | 6.37 | 391,493 | 6.25 | 362,176 | 6.82 | 53,396 | 4.91 |
| Region 2 | 1,160,259 | 9.16 | 510,381 | 8.14 | 523,026 | 9.86 | 126,852 | 11.66 |
| Region 3 | 1,569,242 | 12.39 | 783,971 | 12.51 | 650,355 | 12.26 | 134,916 | 12.41 |
| Region 4 | 2,550,759 | 20.15 | 1,143,897 | 18.25 | 1,179,146 | 22.22 | 227,716 | 20.94 |
| Region 5 | 2,063,800 | 16.30 | 1,109,753 | 17.71 | 813,692 | 15.33 | 140,355 | 12.91 |
| Region 6 | 1,264,201 | 9.99 | 702,843 | 11.22 | 420,067 | 7.92 | 141,291 | 12.99 |
| Region 7 | 710,102 | 5.61 | 344,687 | 5.50 | 330,089 | 6.22 | 35,326 | 3.25 |
| Region 8 | 468,966 | 3.70 | 285,968 | 4.56 | 151,014 | 2.85 | 31,984 | 2.94 |
| Region 9 | 1,521,115 | 12.01 | 697,246 | 11.13 | 671,195 | 12.65 | 152,674 | 14.04 |
| Region 10 | 530,222 | 4.19 | 292,634 | 4.67 | 199,534 | 3.76 | 38,054 | 3.50 |
| Missing/Unknown | 14,978 | 0.12 | 3,740 | 0.06 | 6,320 | 0.12 | 4,918 | 0.45 |
| **Medicare Advantage** |  |  |  |  |  |  |  |  |
| Region 1 | 299,991 | 3.95 | 183,220 | 4.44 | 68,721 | 3.38 | 48,050 | 3.35 |
| Region 2 | 659,422 | 8.68 | 322,677 | 7.81 | 158,791 | 7.80 | 177,954 | 12.42 |
| Region 3 | 687,609 | 9.05 | 405,897 | 9.83 | 140,617 | 6.91 | 141,095 | 9.85 |
| Region 4 | 1,398,447 | 18.41 | 687,666 | 16.65 | 384,073 | 18.87 | 326,708 | 22.81 |
| Region 5 | 1,341,244 | 17.65 | 761,688 | 18.45 | 430,630 | 21.15 | 148,926 | 10.40 |
| Region 6 | 757,143 | 9.97 | 338,536 | 8.20 | 213,501 | 10.49 | 205,106 | 14.32 |
| Region 7 | 271,448 | 3.57 | 165,851 | 4.02 | 81,412 | 4.00 | 24,185 | 1.69 |
| Region 8 | 266,358 | 3.51 | 171,806 | 4.16 | 73,032 | 3.59 | 21,520 | 1.50 |
| Region 9 | 1,505,647 | 19.82 | 861,610 | 20.87 | 375,811 | 18.46 | 268,226 | 18.73 |
| Region 10 | 341,469 | 4.49 | 225,404 | 5.46 | 81,117 | 3.98 | 34,948 | 2.44 |
| Missing/Unknown | 68,519 | 0.90 | 5,057 | 0.12 | 27,906 | 1.37 | 35,556 | 2.48 |
| **Facility/Provider Type** |  |  |  |  |  |  |  |  |
| **Fee-For-Service** |  |  |  |  |  |  |  |  |
| Hospital | 608,776 | 4.81 | 355,801 | 5.68 | 184,171 | 3.47 | 68,804 | 6.33 |
| Office | 3,836,991 | 30.31 | 2,160,151 | 34.47 | 1,088,815 | 20.52 | 588,025 | 54.07 |
| Pharmacy/Mass Immunization Center | 7,632,281 | 60.28 | 3,504,720 | 55.93 | 3,913,588 | 73.75 | 213,973 | 19.68 |
| Skilled Nursing Facility | 205,070 | 1.62 | 59,088 | 0.94 | 46,133 | 0.87 | 99,849 | 9.18 |
| Home Health Agency | 54,483 | 0.43 | 16,806 | 0.27 | 18,171 | 0.34 | 19,506 | 1.79 |
| Other | 310,203 | 2.45 | 162,144 | 2.59 | 52,480 | 0.99 | 95,579 | 8.79 |
| Missing/Unknown | 12,905 | 0.10 | 7,903 | 0.13 | 3,256 | 0.06 | 1,746 | 0.16 |
| **Medicare Advantage** |  |  |  |  |  |  |  |  |
| Hospital | 601,783 | 7.92 | 366,232 | 8.87 | 168,081 | 8.26 | 67,470 | 4.71 |
| Office | 6,194,230 | 81.53 | 3,395,748 | 82.23 | 1,640,826 | 80.61 | 1,157,656 | 80.83 |
| Pharmacy/Mass Immunization Center | 247,023 | 3.25 | 107,791 | 2.61 | 114,614 | 5.63 | 24,618 | 1.72 |
| Skilled Nursing Facility | 113,503 | 1.49 | 35,647 | 0.86 | 23,934 | 1.18 | 53,922 | 3.76 |
| Home Health Agency | 37,356 | 0.49 | 12,041 | 0.29 | 10,768 | 0.53 | 14,547 | 1.02 |
| Other | 386,900 | 5.09 | 202,014 | 4.89 | 73,946 | 3.63 | 110,940 | 7.75 |
| Missing/Unknown | 16,502 | 0.22 | 9,939 | 0.24 | 3,442 | 0.17 | 3,121 | 0.22 |
| **Nursing Home Residency** |  |  |  |  |  |  |  |  |
| **Fee-For-Service** |  |  |  |  |  |  |  |  |
| Nursing Home | 258,722 | 2.04 | 76,366 | 1.22 | 72,911 | 1.37 | 109,445 | 10.06 |
| Non-Nursing Home | 12,401,987 | 97.96 | 6,190,247 | 98.78 | 5,233,703 | 98.63 | 978,037 | 89.94 |
| **Medicare Advantage** |  |  |  |  |  |  |  |  |
| Nursing Home | 128,307 | 1.69 | 42,239 | 1.02 | 28,781 | 1.41 | 57,287 | 4.00 |
| Non-Nursing Home | 7,468,990 | 98.31 | 4,087,173 | 98.98 | 2,006,830 | 98.59 | 1,374,987 | 96.00 |
| **Reason for Medicare Eligibility** |  |  |  |  |  |  |  |  |
| **Fee-For-Service** |  |  |  |  |  |  |  |  |
| Aged without ESRD | 11,688,227 | 92.32 | 5,792,053 | 92.43 | 4,947,713 | 93.24 | 948,461 | 87.22 |
| Aged with ESRD | 2,453 | 0.02 | 1,198 | 0.02 | 534 | 0.01 | 721 | 0.07 |
| Disabled without ESRD | 946,213 | 7.47 | 461,970 | 7.37 | 351,632 | 6.63 | 132,611 | 12.19 |
| Disabled with ESRD | 4,150 | 0.03 | 2,034 | 0.03 | 920 | 0.02 | 1,196 | 0.11 |
| ESRD Only | 19,666 | 0.16 | 9,358 | 0.15 | 5,815 | 0.11 | 4,493 | 0.41 |
| **Medicare Advantage** |  |  |  |  |  |  |  |  |
| Aged without ESRD | 6,525,956 | 85.90 | 3,591,073 | 86.96 | 1,747,902 | 85.87 | 1,186,981 | 82.87 |
| Aged with ESRD | 1,281 | 0.02 | 638 | 0.02 | 201 | 0.01 | 442 | 0.03 |
| Disabled without ESRD | 1,057,456 | 13.92 | 531,301 | 12.87 | 285,211 | 14.01 | 240,944 | 16.82 |
| Disabled with ESRD | 2,493 | 0.03 | 1,224 | 0.03 | 424 | 0.02 | 845 | 0.06 |
| ESRD Only | 10,111 | 0.13 | 5,176 | 0.13 | 1,873 | 0.09 | 3,062 | 0.21 |
| **Medicare-Medicaid Dual Eligibility** |  |  |  |  |  |  |  |  |
| Fee-For-Service |  |  |  |  |  |  |  |  |
| Dual-Eligibility | 1,016,142 | 8.03 | 431,167 | 6.88 | 352,253 | 6.64 | 232,722 | 21.40 |
| Non-Dual-Eligibility | 11,644,567 | 91.97 | 5,835,446 | 93.12 | 4,954,361 | 93.36 | 854,760 | 78.60 |
| Medicare Advantage |  |  |  |  |  |  |  |  |
| Dual-Eligibility | 1,525,629 | 20.08 | 680,331 | 16.48 | 371,003 | 18.23 | 474,295 | 33.11 |
| Non-Dual-Eligibility | 6,071,668 | 79.92 | 3,449,081 | 83.52 | 1,664,608 | 81.77 | 957,979 | 66.89 |
| **Area Deprivation Index** |  |  |  |  |  |  |  |  |
| Fee-For-Service |  |  |  |  |  |  |  |  |
| 1-10(th) | 1,664,327 | 13.15 | 754,394 | 12.04 | 792,014 | 14.93 | 117,919 | 10.84 |
| 11-20 | 1,567,324 | 12.38 | 742,553 | 11.85 | 707,289 | 13.33 | 117,482 | 10.80 |
| 21-30 | 1,636,947 | 12.93 | 806,895 | 12.88 | 716,733 | 13.51 | 113,319 | 10.42 |
| 31-40 | 1,552,555 | 12.26 | 776,106 | 12.38 | 664,653 | 12.52 | 111,796 | 10.28 |
| 41-50 | 1,397,021 | 11.03 | 711,645 | 11.36 | 577,133 | 10.88 | 108,243 | 9.95 |
| 51-60 | 1,231,266 | 9.73 | 629,375 | 10.04 | 497,036 | 9.37 | 104,855 | 9.64 |
| 61-70 | 1,088,332 | 8.60 | 556,441 | 8.88 | 428,563 | 8.08 | 103,328 | 9.50 |
| 71-80 | 936,761 | 7.40 | 481,467 | 7.68 | 352,714 | 6.65 | 102,580 | 9.43 |
| 81-90 | 779,125 | 6.15 | 400,520 | 6.39 | 280,098 | 5.28 | 98,507 | 9.06 |
| 91-100 | 480,784 | 3.80 | 243,548 | 3.89 | 165,771 | 3.12 | 71,465 | 6.57 |
| Missing/Unknown | 326,267 | 2.58 | 163,669 | 2.61 | 124,610 | 2.35 | 37,988 | 3.49 |
| Medicare Advantage |  |  |  |  |  |  |  |  |
| 1-10(th) | 907,090 | 11.94 | 511,620 | 12.39 | 237,861 | 11.68 | 157,609 | 11.00 |
| 11-20 | 850,112 | 11.19 | 501,764 | 12.15 | 205,843 | 10.11 | 142,505 | 9.95 |
| 21-30 | 784,219 | 10.32 | 453,491 | 10.98 | 204,283 | 10.04 | 126,445 | 8.83 |
| 31-40 | 767,536 | 10.10 | 429,586 | 10.40 | 214,146 | 10.52 | 123,804 | 8.64 |
| 41-50 | 755,123 | 9.94 | 419,845 | 10.17 | 210,047 | 10.32 | 125,231 | 8.74 |
| 51-60 | 744,366 | 9.80 | 404,678 | 9.80 | 210,718 | 10.35 | 128,970 | 9.00 |
| 61-70 | 735,973 | 9.69 | 394,594 | 9.56 | 205,841 | 10.11 | 135,538 | 9.46 |
| 71-80 | 699,914 | 9.21 | 365,375 | 8.85 | 192,285 | 9.45 | 142,254 | 9.93 |
| 81-90 | 653,872 | 8.61 | 325,155 | 7.87 | 174,744 | 8.58 | 153,973 | 10.75 |
| 91-100 | 596,688 | 7.85 | 268,040 | 6.49 | 153,202 | 7.53 | 175,446 | 12.25 |
| Missing/Unknown | 102,404 | 1.35 | 55,264 | 1.34 | 26,641 | 1.31 | 20,499 | 1.43 |
| **Concomitant Immunization** |  |  |  |  |  |  |  |  |
| Fee-For-Service |  |  |  |  |  |  |  |  |
| Any Concomitant Immunization | 4,307,970 | 34.03 | 2,208,998 | 35.25 | 1,959,483 | 36.93 | 139,489 | 12.83 |
| COVID-19 Vaccine | 2,756,078 | 21.77 | 1,354,188 | 21.61 | 1,329,088 | 25.05 | 72,802 | 6.69 |
| Pneumococcal Vaccine | 487,013 | 3.85 | 272,009 | 4.34 | 173,040 | 3.26 | 41,964 | 3.86 |
| RSV Vaccine | 883,897 | 6.98 | 484,040 | 7.72 | 381,863 | 7.20 | 17,994 | 1.65 |
| Shingrix Vaccine | 180,982 | 1.43 | 98,761 | 1.58 | 75,492 | 1.42 | 6,729 | 0.62 |
| Medicare Advantage |  |  |  |  |  |  |  |  |
| Any Concomitant Immunization | 169,195 | 2.23 | 88,394 | 2.14 | 62,181 | 3.05 | 18,620 | 1.30 |
| COVID-19 Vaccine | 45,145 | 0.59 | 22,812 | 0.55 | 19,870 | 0.98 | 2,463 | 0.17 |
| Pneumococcal Vaccine | 7,841 | 0.10 | 4,070 | 0.10 | 2,420 | 0.12 | 1,351 | 0.09 |
| RSV Vaccine | 70,009 | 0.92 | 36,008 | 0.87 | 27,314 | 1.34 | 6,687 | 0.47 |
| Shingrix Vaccine | 46,200 | 0.61 | 25,504 | 0.62 | 12,577 | 0.62 | 8,119 | 0.57 |

\*HHS Regions:
Region 1: Connecticut, Maine, Massachusetts, New Hampshire, Rhode Island, and Vermont
Region 2: New Jersey, New York, Puerto Rico, and Virgin Islands
Region 3: Delaware, District of Columbia, Maryland, Pennsylvania, Virginia, and West Virginia
Region 4: Alabama, Florida, Georgia, Kentucky, Mississippi, North Carolina, South Carolina, and Tennessee
Region 5: Illinois, Indiana, Michigan, Minnesota, Ohio, and Wisconsin
Region 6: Arkansas, Louisiana, New Mexico, Oklahoma, and Texas
Region 7: Iowa, Kansas, Missouri, and Nebraska
Region 8: Colorado, Montana, North Dakota, South Dakota, Utah, and Wyoming
Region 9: Arizona, California, Hawaii, Nevada, American Samoa, Commonwealth of the Northern Mariana Islands, Federated State of Micronesia, Guam, Marshall Islands, and Republic of Palau

Table 2 summarizes outcome counts, person-time, and incidence rates by risk and control intervals for all study outcomes. Among both the FFS and MA populations, health outcomes with the highest rates (>100 outcomes per 100,000 person-years) included the stroke outcomes, HS, NHS, NHS/TIA, and TIA.

**Table 2.** Adjusted Outcome Rates Among SCCS Analytic Population, by Risk and Control Intervals and Cohort.

| Health Outcome | Cohort | Risk Interval |  |  | Control Interval |  |  |
| --- | --- | --- | --- | --- | --- | --- | --- |
|  |  | Number of Outcomes | Adjusted Person-Years | Rate (95% CI) | Number of Outcomes | Adjusted Person-Years | Rate (95% CI) |
| Anaphylaxis | *Medicare FFS* |  |  |  |  |  |  |
|  | Any influenza vaccine | 16 | 64,427.77 | 24.83 (14.19, 40.33) | 58 | 449,191.59 | 12.91 (9.80, 16.69) |
|  | High-dose influenza vaccine | * | 31,989.23 | 21.88 (8.80, 45.09) | 29 | 223,077.30 | 13.00 (8.71, 18.67) |
|  | Adjuvanted influenza vaccine | * | 27,000.49 | 29.63 (12.79, 58.38) | 25 | 188,285.44 | 13.28 (8.59, 19.60) |
|  | *Medicare Advantage* |  |  |  |  |  |  |
|  | Any influenza vaccine | * | 36,404.42 | 27.32 (13.10, 50.24) | 45 | 255,034.69 | 17.64 (12.87, 23.61) |
|  | High-dose influenza vaccine | * | 19,952.67 | 25.06 (8.14, 58.48) | 24 | 139,045.20 | 17.26 (11.06, 25.68) |
|  | Adjuvanted influenza vaccine | * | 9,845.25 | 30.47 (6.28, 89.05) | 13 | 68,608.50 | 18.95 (10.09, 32.40) |
| Encephalitis and/or Encephalomyelitis (with ADEM) | *Medicare FFS* |  |  |  |  |  |  |
|  | Any influenza vaccine | 142 | 1,260,542.16 | 11.26 (9.49, 13.28) | 178 | 1,392,918.54 | 12.78 (10.97, 14.80) |
|  | High-dose influenza vaccine | 71 | 627,965.63 | 11.31 (8.83, 14.26) | 88 | 694,521.79 | 12.67 (10.16, 15.61) |
|  | Adjuvanted influenza vaccine | 53 | 531,121.27 | 9.98 (7.47, 13.05) | 71 | 587,524.20 | 12.08 (9.44, 15.24) |
|  | *Medicare Advantage* |  |  |  |  |  |  |
|  | Any influenza vaccine | 52 | 499,499.64 | 10.41 (7.77, 13.65) | 76 | 550,627.98 | 13.80 (10.87, 17.28) |
|  | High-dose influenza vaccine | 28 | 275,407.22 | 10.17 (6.76, 14.69) | 41 | 303,914.78 | 13.49 (9.68, 18.30) |
|  | Adjuvanted influenza vaccine | 14 | 134,688.46 | 10.39 (5.68, 17.44) | 21 | 148,538.02 | 14.14 (8.75, 21.61) |
| Guillain-Barre Syndrome (GBS) | *Medicare FFS* |  |  |  |  |  |  |
|  | Any influenza vaccine | 93 | 1,318,301.10 | 7.05 (5.69, 8.64) | 98 | 1,463,721.80 | 6.70 (5.44, 8.16) |
|  | High-dose influenza vaccine | 46 | 654,858.20 | 7.02 (5.14, 9.37) | 44 | 727,742.90 | 6.05 (4.39, 8.12) |
|  | Adjuvanted influenza vaccine | 40 | 554,986.88 | 7.21 (5.15, 9.81) | 46 | 616,876.13 | 7.46 (5.46, 9.95) |
|  | *Medicare Advantage* |  |  |  |  |  |  |
|  | Any influenza vaccine | 23 | 542,656.15 | 4.24 (2.69, 6.36) | 35 | 600,031.21 | 5.83 (4.06, 8.11) |
|  | High-dose influenza vaccine | 14 | 298,841.49 | 4.68 (2.56, 7.86) | 21 | 330,789.34 | 6.35 (3.93, 9.70) |
|  | Adjuvanted influenza vaccine | * | 146,632.93 | 2.73 (0.74, 6.98) | * | 162,219.38 | 5.55 (2.54, 10.53) |
| Hemorrhagic stroke | *Medicare FFS* |  |  |  |  |  |  |
|  | Any influenza vaccine | 1,949 | 1,306,238.98 | 149.21 (142.66, 155.98) | 2,440 | 1,448,623.87 | 168.44 (161.82, 175.25) |
|  | High-dose influenza vaccine | 1,029 | 649,181.25 | 158.51 (148.97, 168.50) | 1,190 | 720,575.88 | 165.15 (155.89, 174.80) |
|  | Adjuvanted influenza vaccine | 697 | 550,205.08 | 126.68 (117.45, 136.44) | 975 | 610,841.29 | 159.62 (149.75, 169.96) |
|  | *Medicare Advantage* |  |  |  |  |  |  |
|  | Any influenza vaccine | 722 | 585,084.25 | 123.40 (114.56, 132.74) | 828 | 646,597.38 | 128.05 (119.48, 137.08) |
|  | High-dose influenza vaccine | 411 | 321,427.67 | 127.87 (115.80, 140.85) | 440 | 355,581.54 | 123.74 (112.45, 135.86) |
|  | Adjuvanted influenza vaccine | 179 | 158,618.51 | 112.85 (96.92, 130.65) | 227 | 175,374.03 | 129.44 (113.15, 147.42) |
| Non-Hemorrhagic stroke | *Medicare FFS* |  |  |  |  |  |  |
|  | Any influenza vaccine | 5,636 | 1,322,518.91 | 426.16 (415.10, 437.43) | 6,589 | 1,467,906.74 | 448.87 (438.10, 459.84) |
|  | High-dose influenza vaccine | 2,777 | 656,858.87 | 422.77 (407.19, 438.79) | 3,257 | 729,684.54 | 446.36 (431.16, 461.96) |
|  | Adjuvanted influenza vaccine | 2,258 | 556,485.57 | 405.76 (389.20, 422.85) | 2,629 | 618,348.91 | 425.16 (409.07, 441.73) |
|  | *Medicare Advantage* |  |  |  |  |  |  |
|  | Any influenza vaccine | 2,323 | 618,684.23 | 375.47 (360.36, 391.06) | 2,639 | 684,907.98 | 385.31 (370.75, 400.29) |
|  | High-dose influenza vaccine | 1,211 | 339,357.71 | 356.85 (337.03, 377.53) | 1,385 | 376,052.75 | 368.30 (349.16, 388.22) |
|  | Adjuvanted influenza vaccine | 664 | 167,798.09 | 395.71 (366.18, 426.99) | 737 | 185,845.25 | 396.57 (368.45, 426.26) |
| Non-Hemorrhagic stroke or Transient ischemic attack | *Medicare FFS* |  |  |  |  |  |  |
|  | Any influenza vaccine | 8,965 | 1,322,453.20 | 677.91 (663.95, 692.09) | 10,370 | 1,467,984.36 | 706.41 (692.88, 720.14) |
|  | High-dose influenza vaccine | 4,480 | 656,825.04 | 682.07 (662.24, 702.34) | 5,105 | 729,719.17 | 699.58 (680.52, 719.04) |
|  | Adjuvanted influenza vaccine | 3,596 | 556,463.75 | 646.22 (625.27, 667.70) | 4,254 | 618,388.39 | 687.92 (667.40, 708.91) |
|  | *Medicare Advantage* |  |  |  |  |  |  |
|  | Any influenza vaccine | 3,719 | 618,867.23 | 600.94 (581.78, 620.57) | 4,051 | 684,798.70 | 591.56 (573.48, 610.06) |
|  | High-dose influenza vaccine | 2,012 | 339,457.16 | 592.71 (567.09, 619.19) | 2,156 | 375,991.89 | 573.42 (549.47, 598.14) |
|  | Adjuvanted influenza vaccine | 1,020 | 167,848.49 | 607.69 (570.97, 646.16) | 1,143 | 185,814.94 | 615.13 (579.98, 651.85) |
| Transient ischemic attack | *Medicare FFS* |  |  |  |  |  |  |
|  | Any influenza vaccine | 3,521 | 1,331,496.86 | 264.44 (255.78, 273.32) | 3,988 | 1,480,427.42 | 269.38 (261.09, 277.87) |
|  | High-dose influenza vaccine | 1,796 | 661,276.88 | 271.60 (259.18, 284.45) | 1,963 | 735,847.63 | 266.77 (255.10, 278.84) |
|  | Adjuvanted influenza vaccine | 1,422 | 559,889.02 | 253.98 (240.95, 267.53) | 1,695 | 623,213.02 | 271.98 (259.18, 285.24) |
|  | *Medicare Advantage* |  |  |  |  |  |  |
|  | Any influenza vaccine | 1,514 | 636,789.90 | 237.76 (225.93, 250.04) | 1,502 | 705,328.23 | 212.95 (202.32, 224.00) |
|  | High-dose influenza vaccine | 864 | 349,083.97 | 247.50 (231.27, 264.57) | 816 | 387,033.12 | 210.83 (196.62, 225.81) |
|  | Adjuvanted influenza vaccine | 390 | 172,703.86 | 225.82 (203.96, 249.38) | 435 | 191,380.17 | 227.30 (206.44, 249.69) |
| Transverse myelitis | *Medicare FFS* |  |  |  |  |  |  |
|  | Any influenza vaccine | 24 | 1,313,021.07 | 1.83 (1.17, 2.72) | 31 | 1,454,740.41 | 2.13 (1.45, 3.02) |
|  | High-dose influenza vaccine | 11 | 652,261.62 | 1.69 (0.84, 3.02) | 14 | 723,271.16 | 1.94 (1.06, 3.25) |
|  | Adjuvanted influenza vaccine | 11 | 552,759.41 | 1.99 (0.99, 3.56) | 13 | 613,104.50 | 2.12 (1.13, 3.63) |
|  | *Medicare Advantage* |  |  |  |  |  |  |
|  | Any influenza vaccine | * | 296,600.56 | 1.69 (0.55, 3.93) | * | 326,437.00 | 0.31 (0.01, 1.71) |
|  | High-dose influenza vaccine | * | 163,436.45 | 1.84 (0.38, 5.36) | * | 180,066.73 | 0.56 (0.01, 3.09) |
|  | Adjuvanted influenza vaccine | * | 78,696.59 | 1.27 (0.03, 7.08) | -- | 86,648.32 | N/A |

Note: Values <11 are suppressed and denoted as \* and counts equaling 0 are denoted as --

### 3.2 Inferential Results

All health outcomes, except transverse myelitis in the MA population, were sufficiently powered to detect a minimum IRR of 1.2 to 3 for inferential analyses.

#### 3.2.1 Primary Analyses

We did not observe statistically significant elevations of risk for anaphylaxis, encephalitis/encephalomyelitis (with ADEM), GBS, HS, or TM, in SCCS analyses with applicable adjustments for each health outcome (Supplemental Table 6). The results were consistent among both the FFS and MA populations, and within persons receiving any, high-dose, or adjuvanted vaccines (Table 3, Figures 2a and 2b).

**Table 3.**
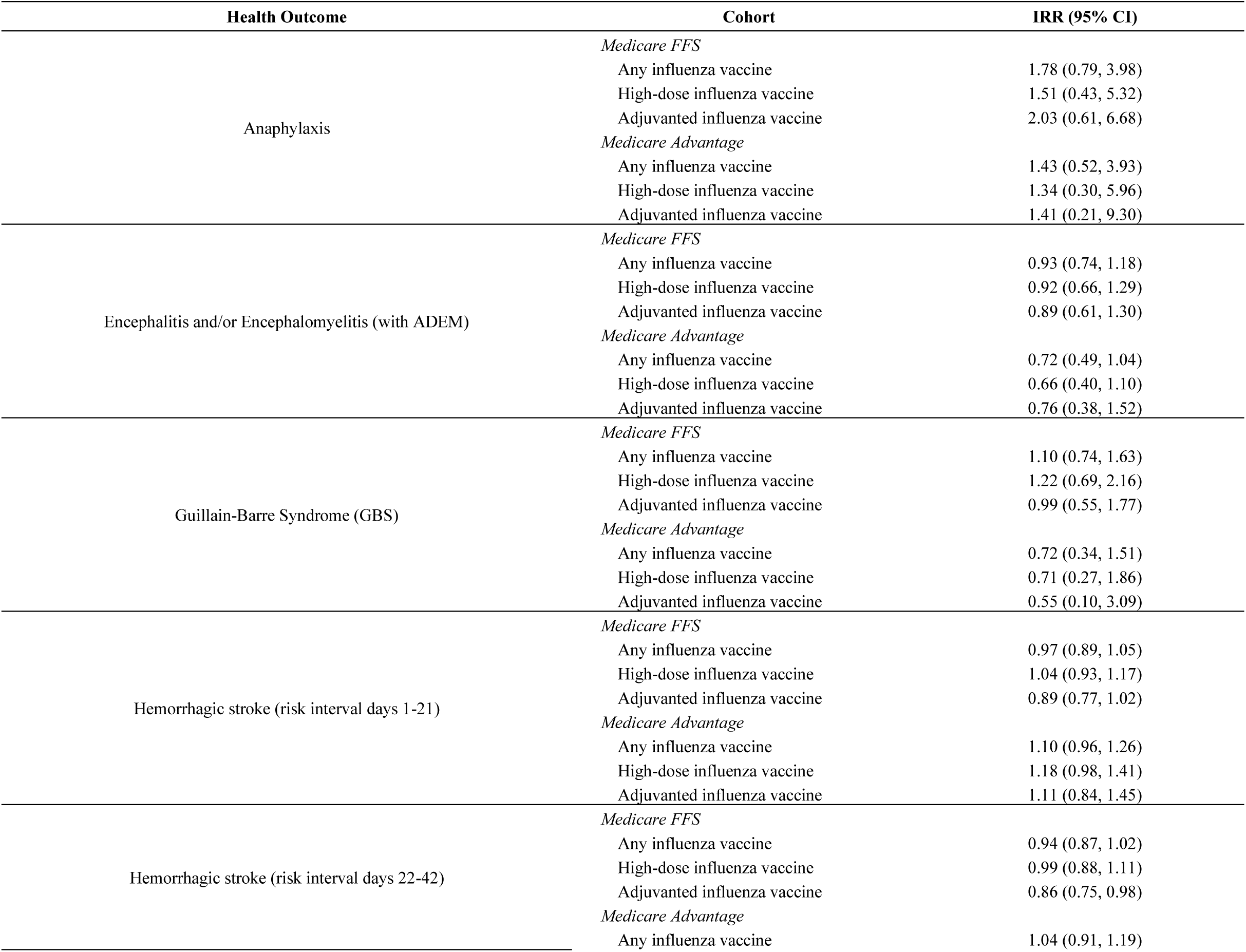
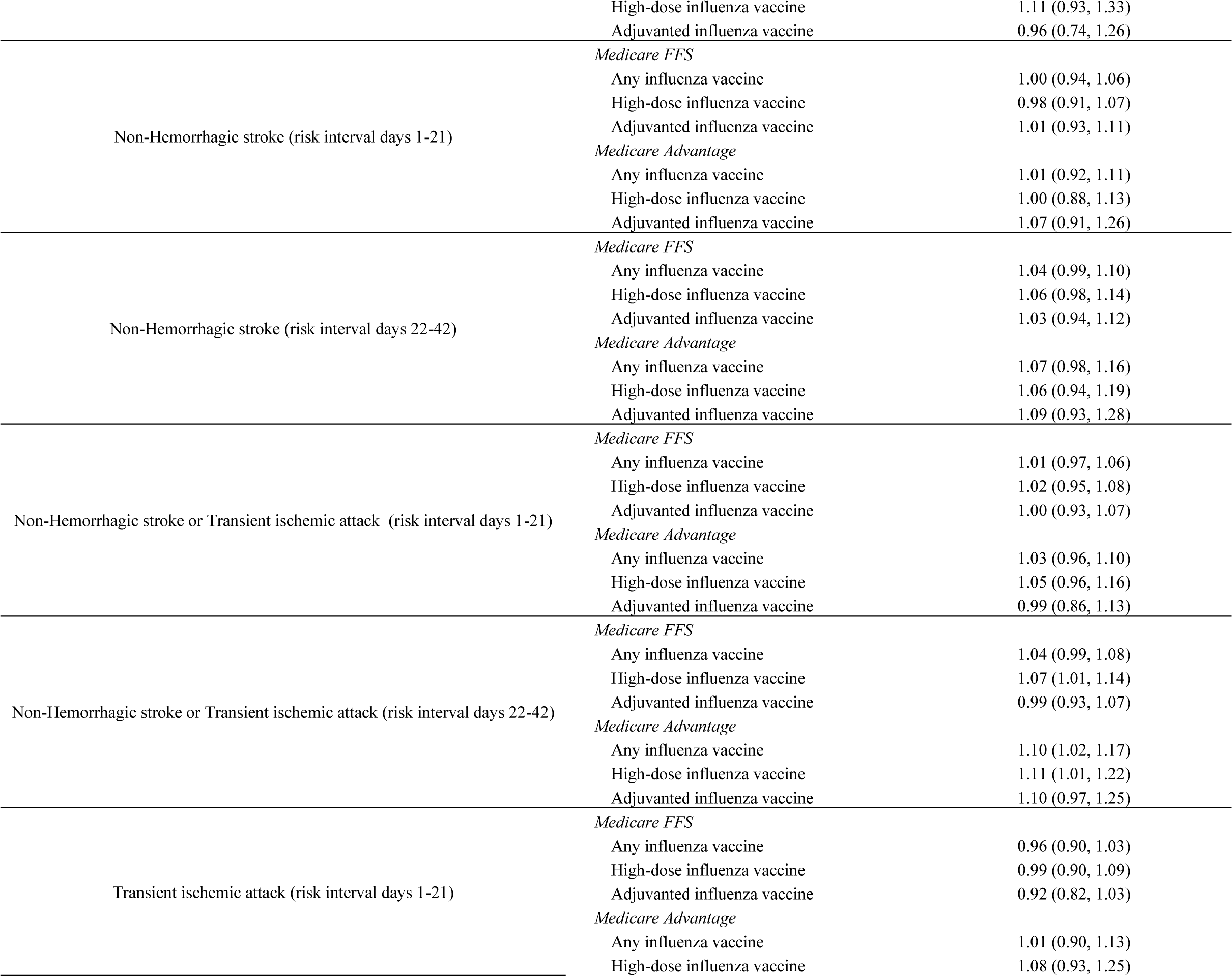
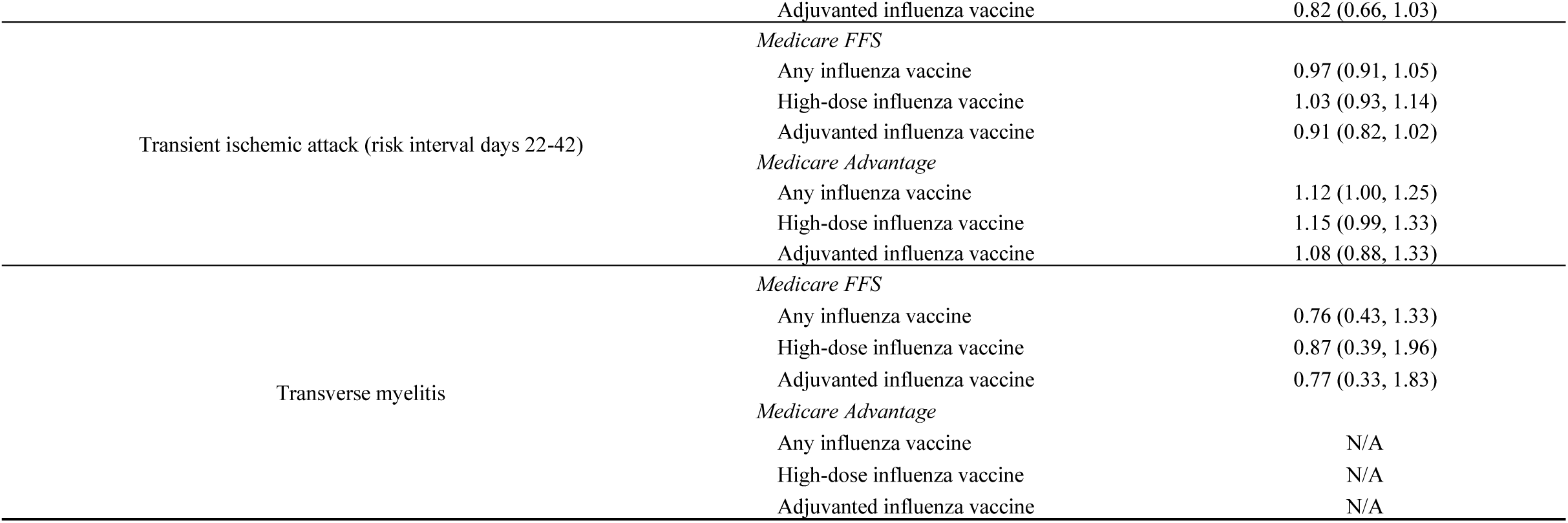
Incidence Risk Ratio (IRR) and 95% CI by CMS Population and Influenza Vaccine Product, Most-Adjusted Analyses.

In the SCCS analyses with seasonality, PPV, and Farrington adjustments, we observed a slightly elevated risk that was statistically significant for the combined outcome of NHS/TIA among both the FFS and MA populations. The small, elevated risk was only observed in the risk window of 22-42 days after high-dose vaccine (FFS and MA). The small risk was also reflected in the MA population that received any influenza vaccine. In the FFS population, the IRR for persons receiving high-dose vaccines was 1.07 (95% CI: 1.01, 1.14). In the MA population, the IRR was 1.11 (95% CI: 1.01, 1.22) following the high-dose vaccine, and 1.10 (95% CI: 1.02, 1.17) following any influenza vaccine. The elevated risk was not observed in persons that received adjuvanted vaccines (FFS IRR: 0.99, 95% CI: 0.93, 1.07, MA IRR: 1.10, 95% CI: 0.97, 1.25) or among FFS beneficiaries that received any influenza vaccine (FFS IRR: 1.04, 95% CI: 0.99, 1.08).

The elevated risk was not consistently observed in the individual NHS or TIA outcomes. For the two NHS and TIA individual outcomes, the only statistically significant elevated risk was observed in the seasonality and PPV-adjusted analysis for TIA in the MA population (IRR: 1.12, 95% CI: 1.00, 1.25). Consistent with the combined NHS/TIA outcome, this risk for TIA was observed only in the 22-42-day risk window, and among persons vaccinated with any influenza vaccine. We also did not observe a significant risk for NHS in either of the FFS or MA populations. Among persons who received a high-dose influenza vaccine, the IRR for NHS was slightly elevated but not statistically significant in the 22-42-day risk window for FFS (IRR: 1.06 95% CI: 0.98, 1.14) and MA (IRR: 1.06, 95% CI: 0.94, 1.19) populations.

#### 3.2.2 Secondary Analyses

We conducted secondary analyses to evaluate the risk of NHS/TIA and TIA following vaccination among those with and without concomitant vaccination. Approximately 27.5% (FFS) and 3.6% (MA) of individuals with NHS/TIA had any concomitant vaccination with the influenza vaccine. The statistically significant risk that we observed in the primary analyses was observed only among individuals without concomitant vaccines (Table 4).

**Table 4.**
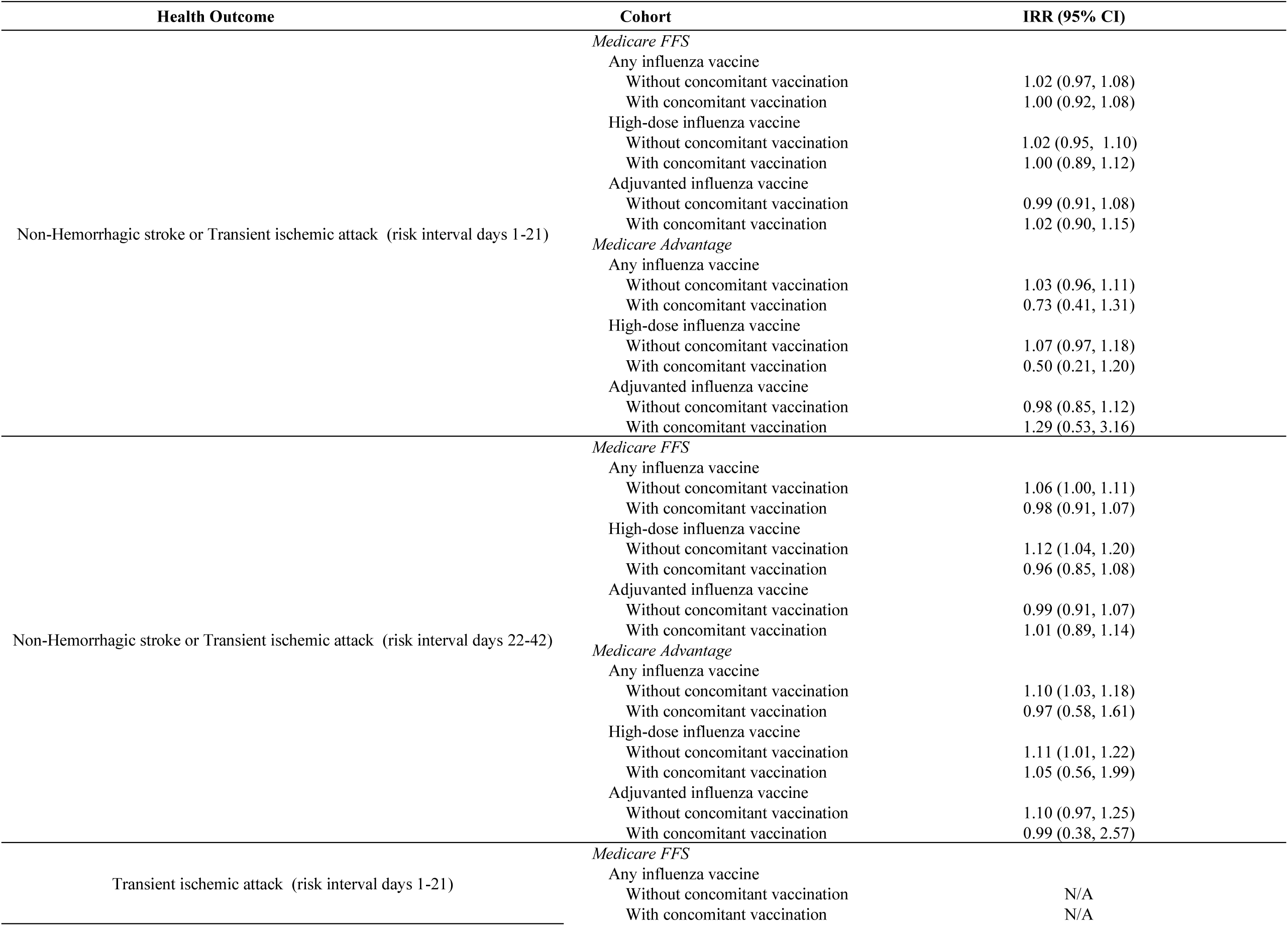
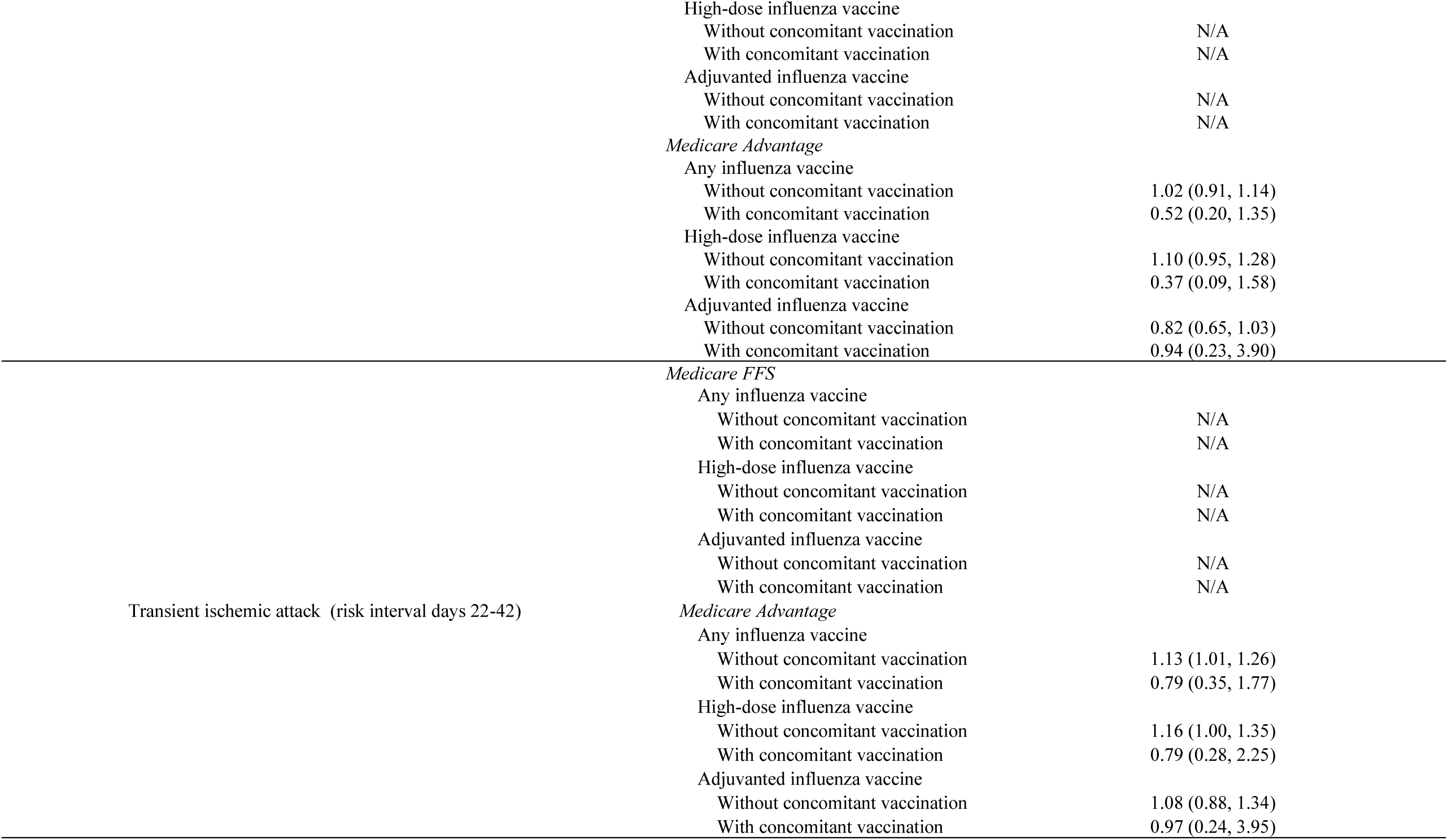
Incidence Risk Ratio (IRR) and 95% CI by CMS Population, Secondary Analyses by Concomitant Vaccination Status, Most-Adjusted Analyses.

For NHS/TIA in the 22-42-day risk window among FFS vaccinees without a concomitant vaccination, we observed a statistically significant risk for high-dose (IRR: 1.12, 95% CI: 1.04, 1.20), and any (IRR: 1.06, 95% CI: 1.00, 1.11) influenza vaccine. Similarly, for NHS/TIA in the 22-42-day risk window among MA vaccinees without a concomitant vaccination, we observed a statistically significant risk for high-dose (IRR: 1.11, 95% CI: 1.01, 1.22) and any (IRR: 1.10, 95% CI: 1.03, 1.18) influenza vaccine. For the individual TIA outcome in the 22-42-day risk window, and among MA vaccinees without a concomitant vaccination, we observed a statistically significant risk for any (IRR: 1.13, 95% CI: 1.01, 1.26) influenza vaccine. We did not observe statistically significant risk among individuals with concomitant vaccines.

#### 3.2.3 Sensitivity Analyses

In our sensitivity analyses assessing a 14-day washout period between risk and control intervals, results remained consistent for all outcomes for which we did not observe significantly elevated risk in the primary analyses (Table 5). The statistically significant IRR for NHS/TIA in the 22-42-day risk window following a high-dose vaccine that was observed in the FFS (IRR: 1.06, 95% CI: 0.99, 1.14) and MA (IRR: 1.09, 95% CI: 0.98, 1.20) populations were no longer significant in the washout analysis. However, a statistically significant elevation in risk for NHS/TIA among the MA population persisted for any (IRR: 1.08, 95% CI: 1.00, 1.16) influenza vaccine (Table 5). Finally, the small, elevated risk for the individual TIA outcome in the 22-42-day risk window was no longer significant among the MA population that received any influenza vaccine (IRR: 1.09, 95% CI: 0.97, 1.23).

**Table 5.**
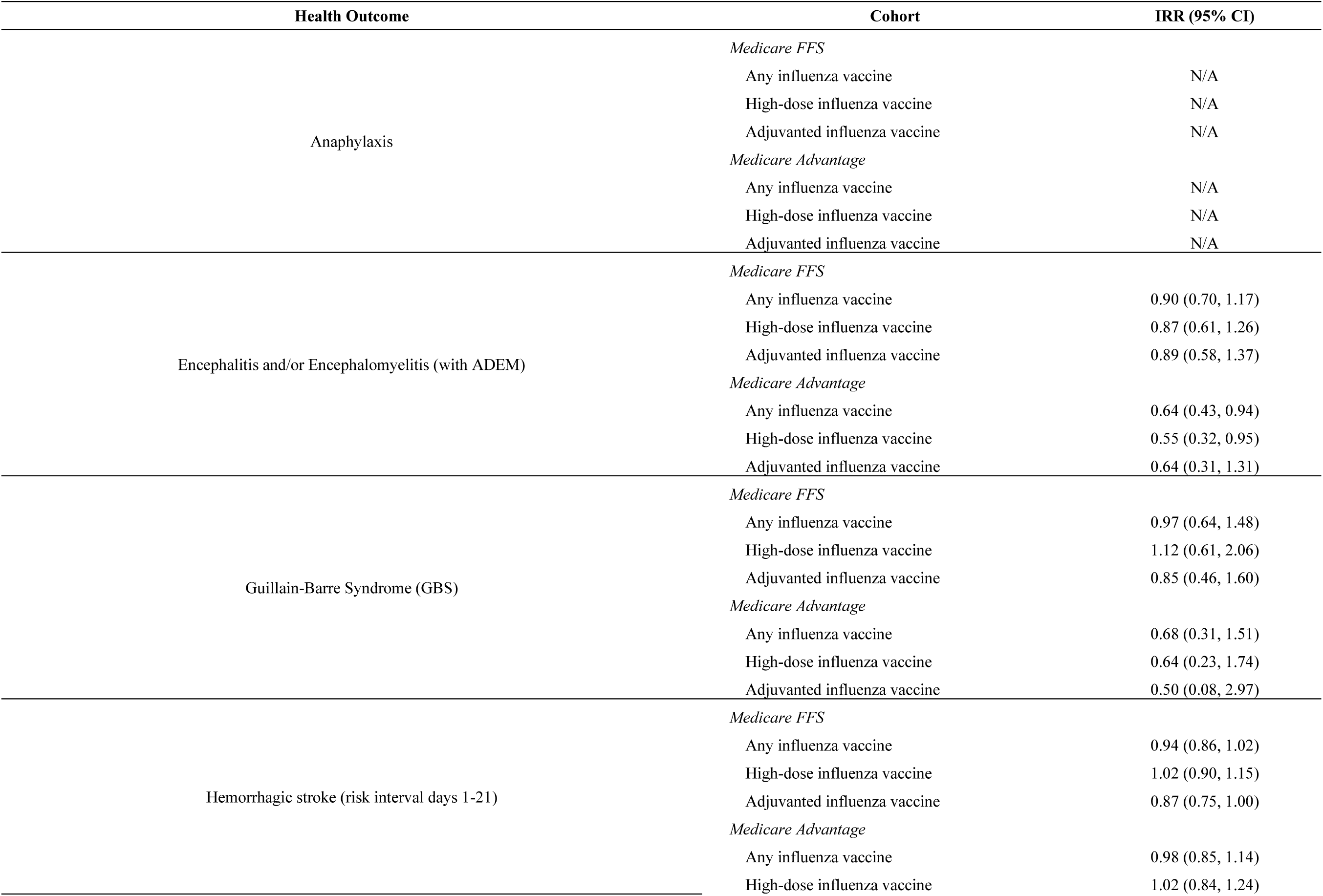
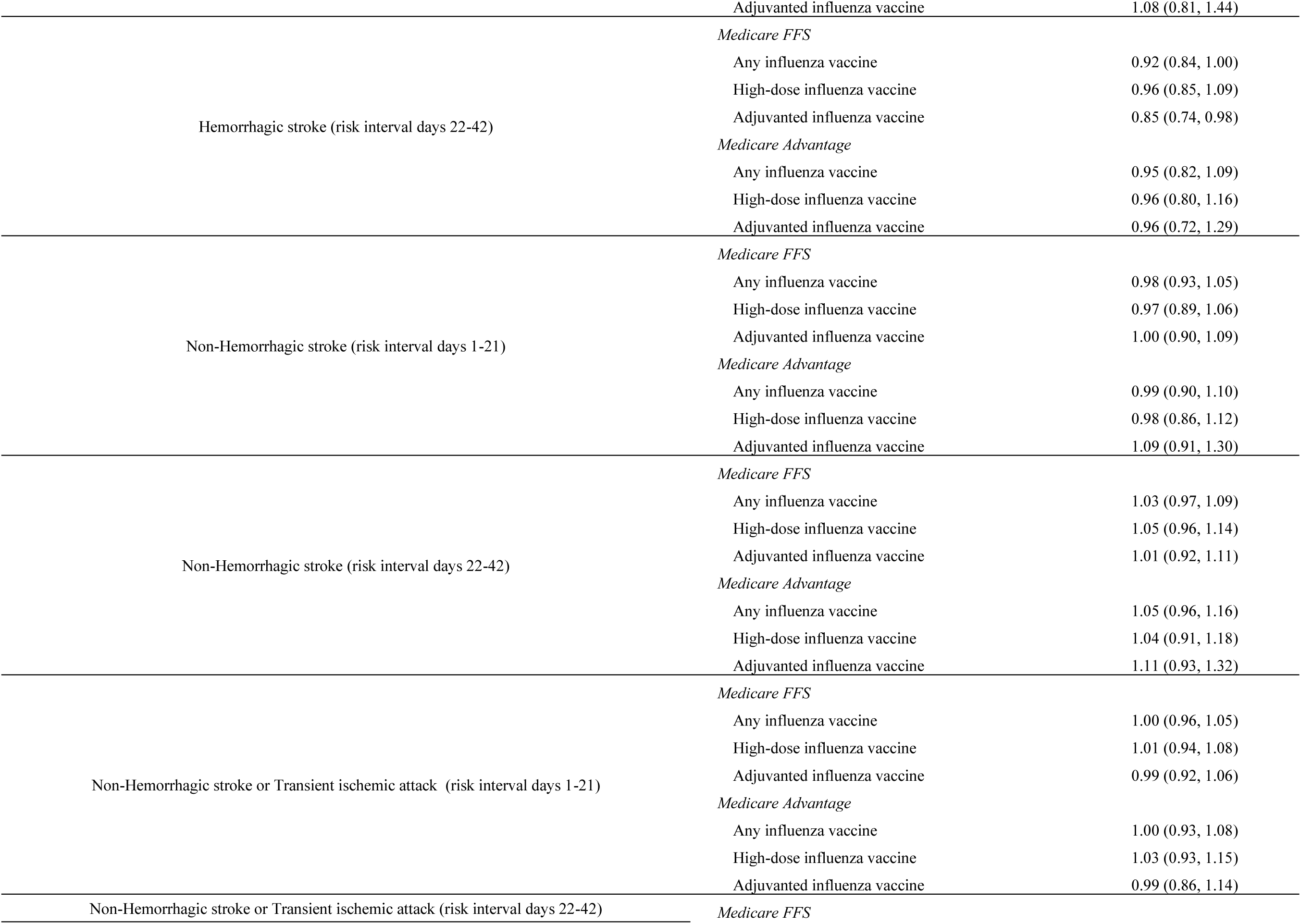
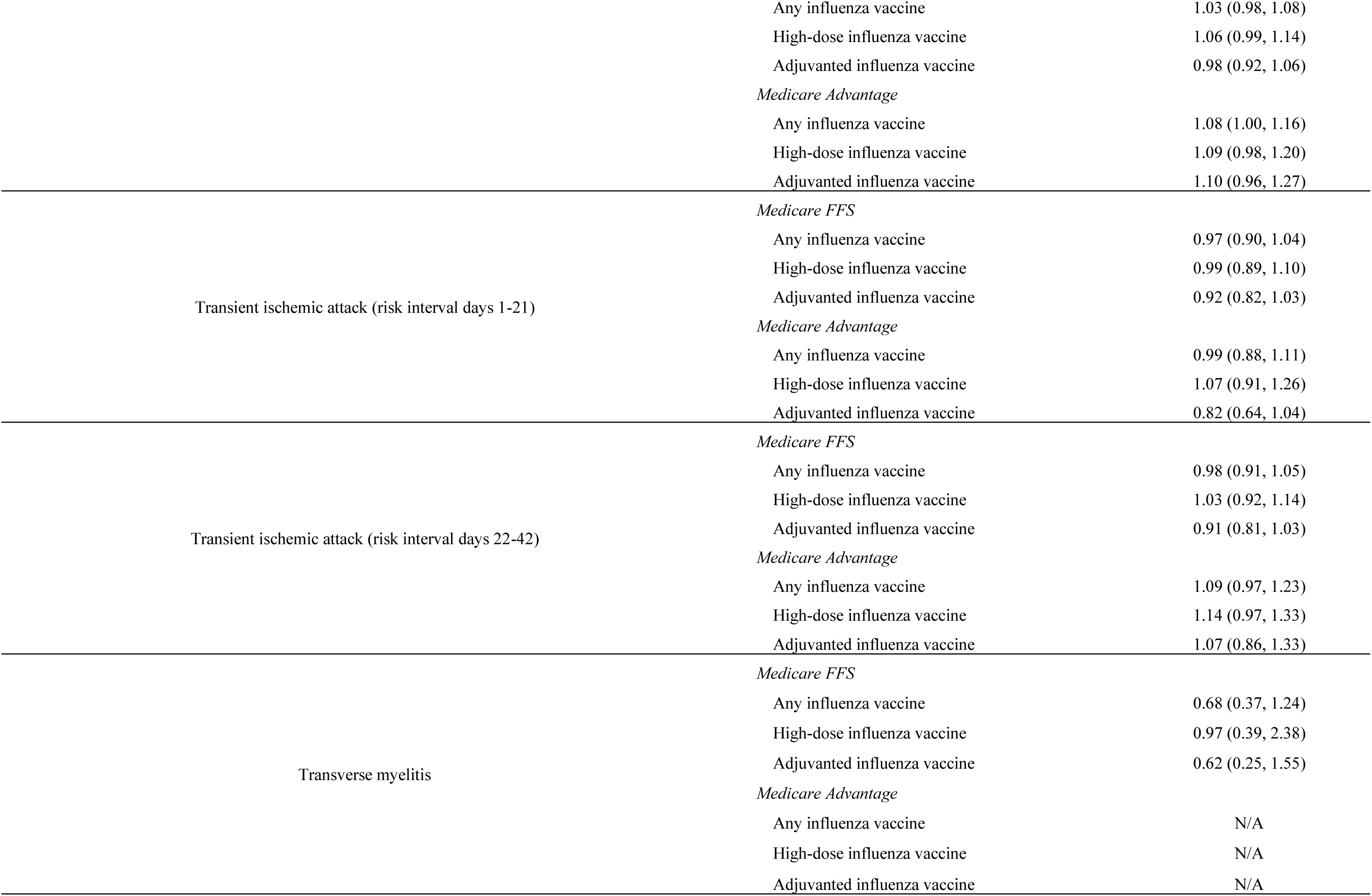
Incidence Risk Ratio (IRR) and 95% CI by CMS Population, Washout Period Sensitivity Analyses, Most-Adjusted Analyses.

We did not observe statistically significant elevations in risk for any health outcome in the full planned observation period sensitivity analyses for the FFS and MA populations (Table 6). In the FFS population, the IRR for NHS/TIA (22-42-day risk window) was 1.03 (95% CI: 0.97, 1.09) for the high-dose vaccine and 1.00 (95% CI: 0.96, 1.04) for any influenza vaccine. The corresponding IRR in the MA population was 1.07 (95% CI: 0.98, 1.17) for the high-dose vaccine, and 1.06 (95% CI: 0.99, 1.13) for any influenza vaccine.

**Table 6.**
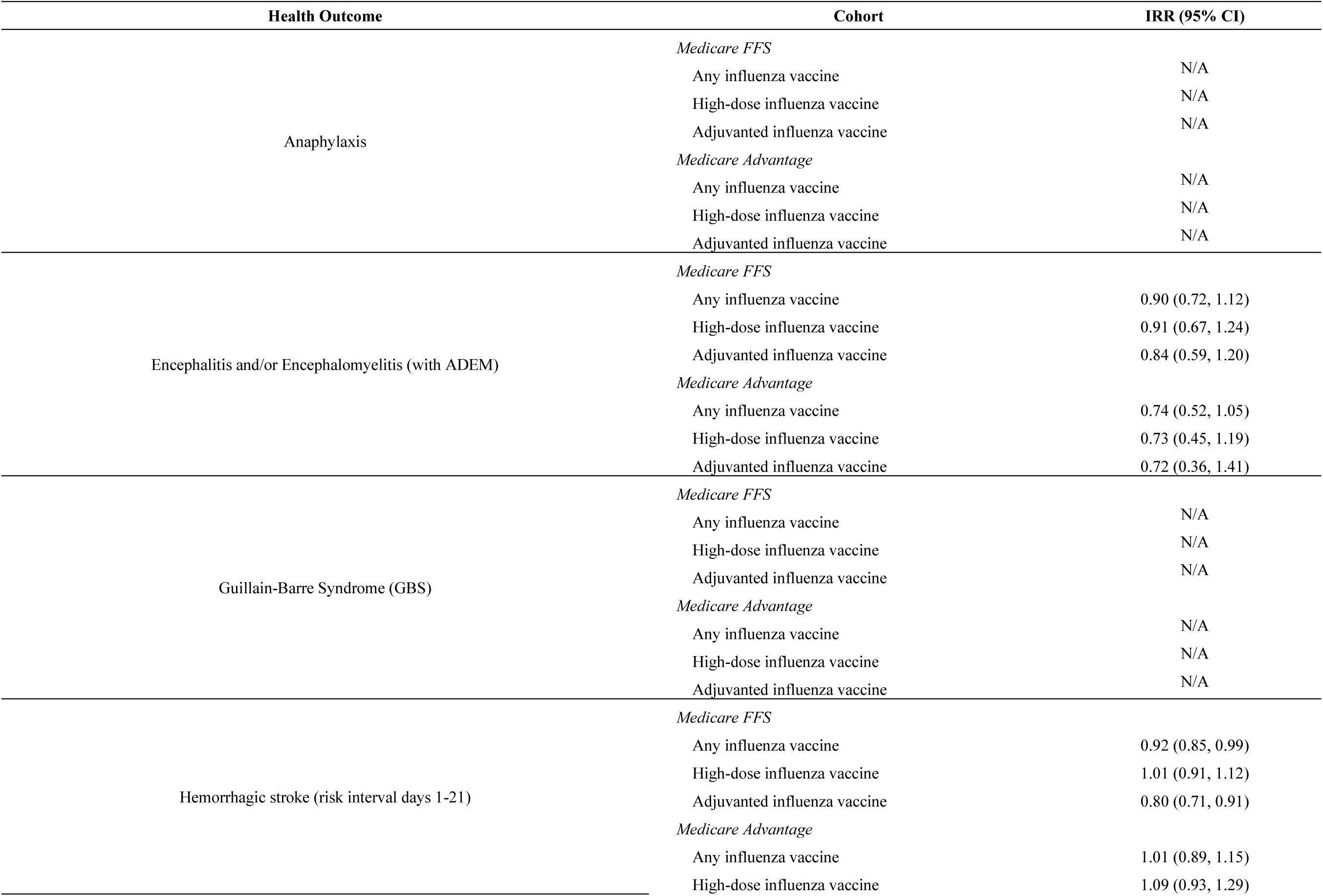
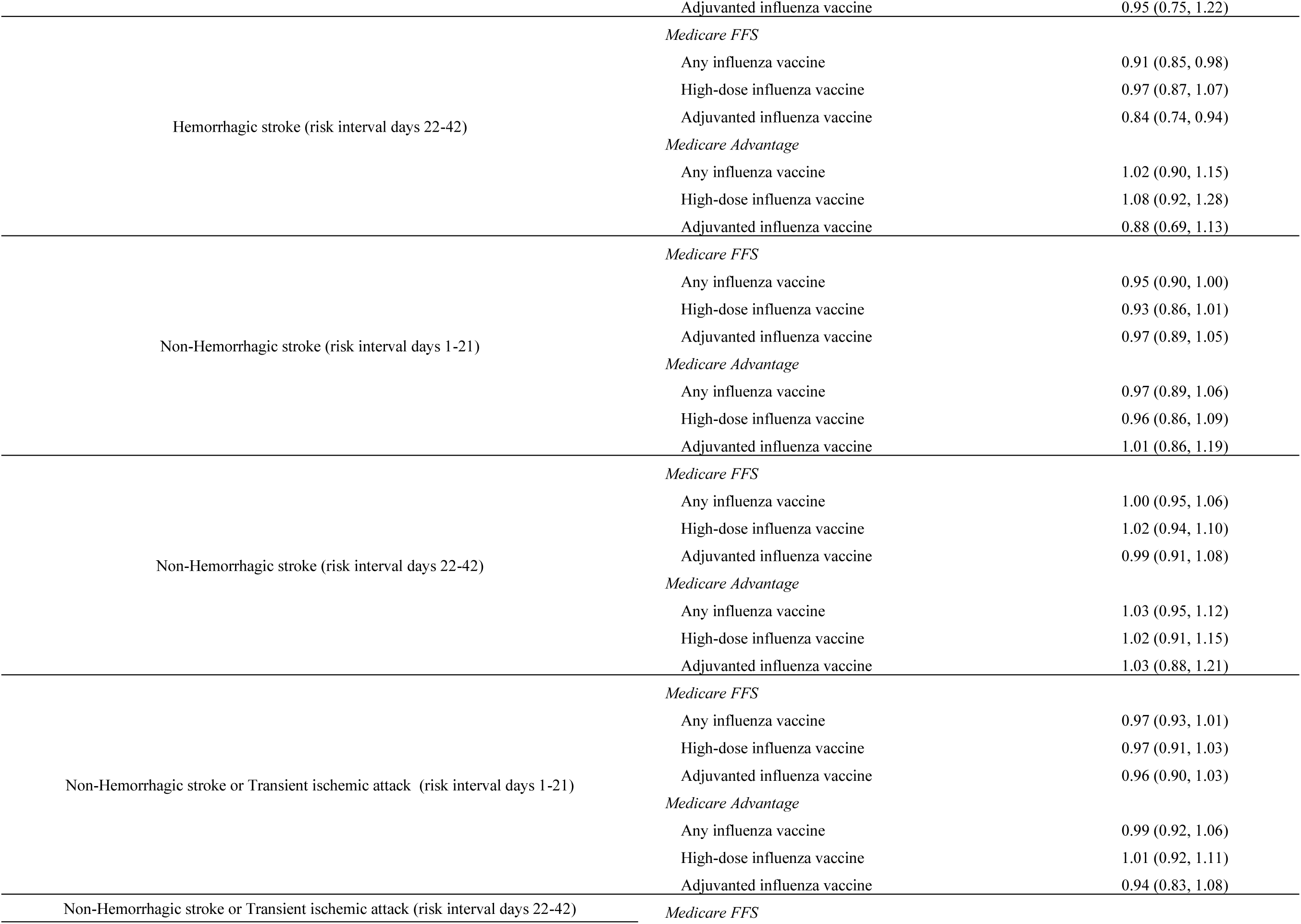
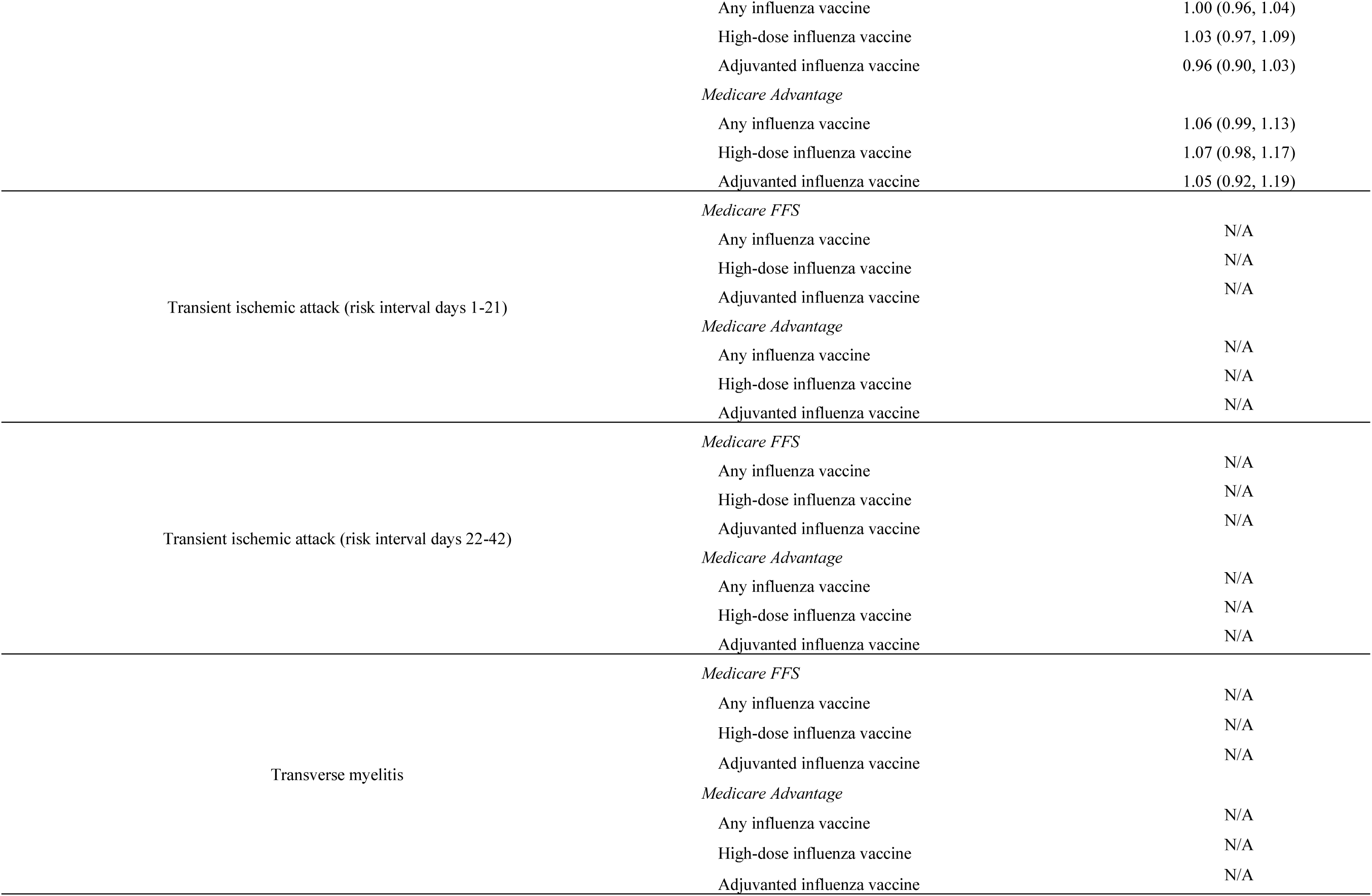
Incidence Risk Ratio (IRR) and 95% CI by CMS Population, Full Planned Observation Period Sensitivity Analyses, Most-Adjusted Analyses.

## DISCUSSION

In this study of approximately 20 million Medicare beneficiaries aged 65 years and older that received an annual influenza vaccine, small but statistically significant elevations in risk for NHS/TIA and TIA in the 22–42-day risk interval were observed. However, no statistically significant elevations in risk were observed for NHS/TIA and TIA in our sensitivity analyses for Medicare FFS population. In our washout sensitivity analyses, a small significant elevation in risk for TIA (22–42-day risk interval) persisted among MA beneficiaries who had received any influenza vaccine. There were no statistically significant elevations of risk for anaphylaxis, encephalitis/encephalomyelitis (with ADEM), GBS, HS, or TM, in adjusted SCCS analyses in the Medicare FFS or MA populations. In our concomitant vaccination analysis for NHS/TIA and TIA, we observed statistically significant elevations in risk among individuals who did not receive a concomitant vaccination.

While influenza vaccines are generally considered safe based on studies conducted from previous seasons, some studies have reported an increased risk of health outcomes following influenza vaccination.^(6, 7, 15, 16)^ In a self-controlled case series study of several health outcomes following influenza vaccination among adults aged 65 years and older during three influenza seasons (2016–2017, 2017–2018 and 2018–2019), an increased stroke risk was observed following high-dose or adjuvanted influenza vaccination in the primary and age subgroup analyses, but these findings were not consistent across the three respiratory seasons.^(12)^ During the 2023-2024 respiratory season, we identified a small yet statistically significant increase in risk; in all IRR estimates for NHS/TIA and TIA, the magnitude of the IRR was less than 1.2 which may be due to difficulty in diagnosing TIA.^(17)^

Our study has several strengths. First, our study was conducted in a large administrative claims database utilizing both FFS and MA beneficiaries, representing the majority of U.S adults aged 65 years and older. With more than 20 million influenza vaccine doses included in the study, our study was well-powered to identify rare health outcomes following vaccination. Second, we used a self-controlled design comparing the rate of each outcome in risk interval to the rate in control interval within each individual which enables control for time-invariant confounding. This is particularly important in our population that may experience health concerns that increase the risk for health outcomes under evaluation. Third, we incorporated adjustments to several analyses including accounting for seasonality and event-dependent observation time. Finally, when the PPV was available from prior medical record review, a PPV multiple imputation-based quantitative bias analysis helped to account for uncertainty in our IRR and AR estimates due to potential miscoding in claims data.

Our study also has several limitations for consideration. First, outcome misclassification is a concern as cases were identified using administrative claims data. However, we implemented PPV-adjusted analyses to partially address this potential misclassification. Second, the post-vaccination risk and control intervals for the health outcomes may have been mis-specified, but we expect that the washout sensitivity analyses partially accounted for this limitation. The risk and control intervals were determined based on clinician consultation, but it may be difficult to determine when the rates of adverse events post-vaccination return to baseline. Third, this design does not account for time-varying confounders and does not adjust for changes over time among vaccinated individuals. However, we expect that the impact of time-varying confounding is minimal since the total follow-up time for any individual was 90 days. Finally, despite the large administrative claims database used for this study, sample size may have impacted secondary concomitant vaccination analyses or subgroup analyses when evaluating rare outcomes.

Inconsistent results in the composite NHS/TIA and individual outcomes (NHS or TIA) were potentially due to insufficient statistical power in the subpopulations. With the two individual outcomes, the smaller sample size lowered statistical power to accurately detect small IRRs.

## Conclusion

Through an FDA-CMS partnership, this study evaluated the safety of the 2023–2024 seasonal influenza vaccines among U.S. adults aged 65 years and older and found a small yet statistically significant increase in risk of the composite outcome of NHS/TIA and TIA following influenza vaccination. There were no statistically significant elevations in incidence rates of encephalitis/encephalomyelitis, GBS, or transverse myelitis observed to be associated with influenza vaccines. The potential risk of NHS/TIA and TIA following vaccination must be carefully considered with known benefits of influenza vaccination. Based on the results from this study, FDA believes the benefits of seasonal influenza vaccination continue to outweigh the risks.

## Data Availability

All data produced in the present study are available upon reasonable request to the authors.

## Acknowledgments

We thank Rose Do for her support in providing clinical insight to study findings, and Bradley Lufkin for his support with manuscript drafting and review. We appreciate review and feedback provided by Dr. Henry Zhang and Dr. Carla Zelaya on the manuscript. We thank Ms. Krista Fekecs for project support.

## Footnotes

a Concomitant vaccines for Medicare beneficiaries included COVID-19, PCV/PPSV, RSV, and Shingrix.

b CFR calculated in the FFS SCCS study population was also applied in analyses for the MA population

