## Supplemental Material for "Safety Monitoring of Health Outcomes following Influenza Vaccination during the 2023-2024 Season among U.S. Medicare Beneficiaries Aged 65 Years and Older"

**Supplementary Table 1. 2023-2024 Influenza Vaccines and Recommended Age Groups, Medicare Population**

| Name | Age Group |
|---|---|
| Fluad Quadrivalent (2023-2024) | 65 years and older |
| Fluzone High-Dose Quadrivalent (2023-2024) | 65 years and older |
| Flucelvax Quadrivalent (2023-2024) | 6 months and older |
| Flublok Quadrivalent (2023-2024) | 18 years and older |
| Fluarix Quadrivalent (2023-2024) | 6 months and older |
| Flulaval Quadrivalent (2023-2024) | 6 months and older |
| Fluzone Quadrivalent (2023-2024) | 6 months and older |
| Afluria Quadrivalent (2023-2024) | 6 months and older |

**Supplementary Table 2. Influenza Vaccine (2023-2024 Formula) Product and Code List, Medicare Population**

| Manufacturer | Name | Vaccine Group | Code Type | Code |
|---|---|---|---|---|
| Seqirus | Fluad Quadrivalent (2023-2024) | Adjuvanted | CPT | 90688 |
| Seqirus | Fluad Quadrivalent (2023-2024) | Adjuvanted | CPT | 90694 |
| Seqirus | Fluad Quadrivalent (2023-2024) | Adjuvanted | NDC | 70461-0123-03 |
| Seqirus | Fluad Quadrivalent (2023-2024) | Adjuvanted | NDC | 70461-0123-04 |
| Sanofi Pasteur | Fluzone High-Dose Quadrivalent (2023-2024) | High-Dose | CPT | 90662 |
| Sanofi Pasteur | Fluzone High-Dose Quadrivalent (2023-2024) | High-Dose | NDC | 49281-0123-65 |
| Sanofi Pasteur | Fluzone High-Dose Quadrivalent (2023-2024) | High-Dose | NDC | 49281-0123-88 |
| Seqirus | Flucelvax Quadrivalent (2023-2024) | Other | CPT | 90674 |
| Seqirus | Flucelvax Quadrivalent (2023-2024) | Other | NDC | 70461-0323-03 |
| Seqirus | Flucelvax Quadrivalent (2023-2024) | Other | NDC | 70461-0323-04 |
| Seqirus | Flucelvax Quadrivalent (2023-2024) | Other | NDC | 70461-0423-10 |
| Seqirus | Flucelvax Quadrivalent (2023-2024) | Other | NDC | 70461-0423-11 |
| Sanofi Pasteur | Flublok Quadrivalent (2023-2024) | Other | CPT | 90682 |
| Sanofi Pasteur | Flublok Quadrivalent (2023-2024) | Other | NDC | 49281-0723-88 |
| Sanofi Pasteur | Flublok Quadrivalent (2023-2024) | Other | NDC | 49281-0723-10 |
| GlaxoSmithKline Biologicals | Fluarix Quadrivalent (2023-2024) | Other | CPT | 90686 |
| GlaxoSmithKline Biologicals | Fluarix Quadrivalent (2023-2024) | Other | NDC | 58160-0909-52 |
| GlaxoSmithKline Biologicals | Fluarix Quadrivalent (2023-2024) | Other | NDC | 58160-0909-41 |
| GlaxoSmithKline Biologicals | Flulaval Quadrivalent (2023-2024) | Other | CPT | 90686 |
| GlaxoSmithKline Biologicals | Flulaval Quadrivalent (2023-2024) | Other | NDC | 19515-0814-52 |
| GlaxoSmithKline Biologicals | Flulaval Quadrivalent (2023-2024) | Other | NDC | 19515-0814-41 |
| Sanofi Pasteur | Fluzone Quadrivalent (2023-2024) | Other | CPT | 90686 |
| Sanofi Pasteur | Fluzone Quadrivalent (2023-2024) | Other | NDC | 49281-0423-50 |
| Sanofi Pasteur | Fluzone Quadrivalent (2023-2024) | Other | NDC | 49281-0423-88 |
| Seqirus | Afluria Quadrivalent (2023-2024) | Other | CPT | 90686 |
| Seqirus | Afluria Quadrivalent (2023-2024) | Other | CPT | 90688 |
| Seqirus | Afluria Quadrivalent (2023-2024) | Other | NDC | 33332-0323-03 |
| Seqirus | Afluria Quadrivalent (2023-2024) | Other | NDC | 33332-0323-04 |
| Sanofi Pasteur | Fluzone Quadrivalent (2023-2024) | Other | CPT | 90688 |
| Sanofi Pasteur | Fluzone Quadrivalent (2023-2024) | Other | NDC | 49281-0639-78 |
| Sanofi Pasteur | Fluzone Quadrivalent (2023-2024) | Other | NDC | 49281-0639-15 |
| Seqirus | Flucelvax Quadrivalent (2023-2024) | Other | CPT | 90756 |
| Seqirus | Flucelvax Quadrivalent (2023-2024) | Other | NDC | 70461-0323-03 |

| | | | | |
|---|---|---|---|---|
| Seqirus | Flucelvax Quadrivalent (2023-2024) | Other | NDC | 70461-0323-04 |
| AstaZeneca | FluMist Quadrivalent (2023-2024) | Other | CPT | 90672 |
| AstaZeneca | FluMist Quadrivalent (2023-2024) | Other | NDC | 66019-0310-10 |
| AstaZeneca | FluMist Quadrivalent (2023-2024) | Other | NDC | 66019-0310-01 |

**Supplementary Table 3. Outcomes, Care Settings, Risk Intervals, Control Intervals, Clean Windows, and Codes used in Surveillance**

| Outcomes | Care Setting | Clean Window* | Risk Interval** | Control Interval*** | Code List |
|---|---|---|---|---|---|
| Anaphylaxis | IP, OP-ED | 30 days | 0-1 day | 2-16 days | T8052XAM T782XXA |
| Encephalitis/Encephalomyelitis /ADEM | IP | 183 days | 1-42 days | 43-90 days | G0402 G0400 G0481 G0490 G053 G0481 G0490 G053 |
| Transverse myelitis† | IP, OP-ED | 365 days | 1-42 day | 43-90 days | G373 |
| Non-hemorrhagic stroke/Transient Ischemic Attack† | IP, OP-ED (TIA only) | 365 days | 1-21 days<br>22-42 days | 43-90 days | I6300 I63011 I63012 I63013 I63019 I6302 I63031 I63032 I63033 I63039 I6309 I6310 I63111 I63112 I63113 I63119 I6312 I63131 I63132 I63133 I63139 I6319 I6320 I63211 I63212 I63213 I63219 I6322 I63231 I63232 I63233 I63239 I6329 I6330 I63311 I63312 I63313 I63319 I63321 I63322 I63323 I63329 I63331 I63332 I63333 I63339 I63341 I63342 I63343 I63349 I6339 I6340 I63411 I63412 I63413 I63419 I63421 I63422 I63423 I63429 I63431 I63432 I63433 I63439 I63441 I63442 I63443 I63449 I6349 I6350 I63511 I63512 I63513 I63519 I63521 I63522 I63523 I63529 I63531 I63532 I63533 I63539 I63541 I63542 I63543 I63549 I6359 I636 I6381 I6389 I639 G458 G459 |
| Non-hemorrhagic stroke† | IP | 365 days | 1-21 days<br>22-42 days | 43-90 days | I6300 I63011 I63012 I63013 I63019 I6302 I63031 I63032 I63033 I63039 I6309 I6310 I63111 I63112 I63113 I63119 I6312 I63131 I63132 I63133 I63139 I6319 I6320 I63211 I63212 I63213 I63219 I6322 I63231 I63232 I63233 I63239 I6329 I6330 I63311 I63312 I63313 I63319 I63321 I63322 I63323 I63329 I63331 I63332 I63333 I63339 I63341 I63342 I63343 I63349 I6339 I6340 I63411 I63412 I63413 I63419 I63421 I63422 I63423 I63429 I63431 I63432 I63433 I63439 I63441 I63442 I63443 I63449 I6349 I6350 I63511 I63512 I63513 I63519 I63521 I63522 I63523 I63529 I63531 I63532 I63533 I63539 I63541 I63542 I63543 I63549 I6359 I636 I6381 I6389 I639 |
| Hemorrhagic Stroke† | IP | 365 days | 1-21 days<br>22-42 days | 43-90 days | I610 I611 I612 I613 I614 I615 I616 I618 I619 I6200 I6201 I6202 I629 |
| Transient Ischemic Attack† | IP, OP-ED | 365 days | 1-21 days<br>22-42 days | 43-90 days | G458 G459 |
| Guillain-Barré syndrome | IP- primary position only | 365 days | 1-42 days | 43-90 days | G610 |

* Clean Window is used to define incident outcomes where an individual enters the study cohort only if the outcome of interest did not occur during that interval prior to the observed outcome. References for the duration of the clean window were not available in existing literature and are instead based on clinician input.

** Risk intervals are determined based on clinical guidance and literature review and are defined as an interval during which excess risk is hypothesized following influenza vaccination.

*** Control interval is defined as all follow-up time during the observation period that is not in the risk interval or in a washout period.

† The algorithm used to identify incident outcomes are complex and are referenced in Supplementary Table 4

**Supplementary Table 4. Adjustments and Exclusions for HS, NHS, NHS/TIA, and TIA**

| Outcome | Adjust onset date if observed in the 1 day prior to outcome (in all settings) | Exclusions for Prevalence (in all settings) | Exclusions – other known causes (in all settings) |
|---|---|---|---|
| Hemorrhagic Stroke (HS) | I63.9, R51*, R47*, R29.810, R53.1, R42*, R41.82, R40.4, H53.13*, H53.9, G81.9* | If occurs in the 365 window prior to outcome<br>I69*, Z86.73 | If in last 30 days prior to outcome<br>U07.1<br>If in last 1 day prior to outcome<br>S06*<br>If same day as outcome<br>S06*, Physical Trauma Code |
| Non-Hemorrhagic Stroke (NHS) | Z92.82, R51*, R47*, R29.810, R53.1, R42, R41.82, R40.4, G81.9*, H53.9, H53.13* | If occurs in the 365 window prior to outcome<br>I69*, Z86.73, I48*, D57*, D68.5* | If in last 30 days prior to outcome<br>U07.1<br>If in last 28 days prior to outcome<br>I21*<br>If in last 1 day prior to outcome<br>S15*, 174*<br>In same day as outcome<br>S15*, I74*, Physical Trauma Code |
| Non-hemorrhagic stroke or Transient Ischemic Attacks (NHS/TIA) | Z92.82, R51*, R47*, R29.810, R53.1, R42, R41.82, R40.4, G81.9*, H53.9, H53.13* | If occurs in the 365 window prior to outcome<br>I69*, Z86.73,<br>I48*, D57*, D68.5* | If in last 30 days prior to outcome<br>U07.1<br>If in last 28 days prior to outcome<br>I21*<br>If in last 1 day prior to outcome<br>S15*, 174*<br>In same day as outcome<br>S15*, I74*, Physical Trauma Code |
| Transient Ischemic Attacks (TIA) | R51*, R47*, R29.810, R53.1, R42, R41.82, R40.4, G81.9*, H53.9, H53.13* | If occurs in the 365 window prior to outcome<br>I69*, Z86.73, I48*, D57*, D68.5* | If in last 30 days prior to outcome<br>U07.1<br>If in last 28 days prior to outcome<br>I21*<br>If in last 1 day prior to outcome<br>S15*<br>In same day as outcome<br>S15*, Physical Trauma Code |

**Supplementary Table 5. Outcome Positive Predictive Value (PPV) Estimates from Prior MRR**

| Outcome | PPV (95% CI) |
|---|---|
| Anaphylaxis (IP, OP-ED) | 66% (56%, 76%) |
| Guillain-Barré syndrome (IP, primary diagnosis position) | 71% (63%, 79%) |
| Non-hemorrhagic stroke (IP) | 80% (71%, 87%) |
| Transient ischemic attack (IP, OP-ED) | 77% (65%, 86%) |
| Non-hemorrhagic stroke (IP) or Transient ischemic attack (IP, OP-ED)* | 79% (70%, 88%) |

* PPV value for this combined outcome is estimated by the weighted average of PPV values of the individual NHS and TIA outcomes

**Supplementary Table 6. Most-Adjusted SCCS Analyses for Each Health Outcome**

| Health Outcome | Most-Adjusted SCCS Analysis |
|---|---|
| Anaphylaxis | Seasonality and PPV adjusted |
| Encephalitis and/or Encephalomyelitis (with ADEM) | Seasonality and Farrington adjusted |
| Guillain-Barre Syndrome (GBS) | Seasonality and PPV adjusted |
| Hemorrhagic stroke (risk interval days 1-21) | Seasonality and Farrington adjusted |
| Hemorrhagic stroke (risk interval days 22-42) | Seasonality and Farrington adjusted |
| Non-Hemorrhagic stroke (risk interval days 1-21) | Seasonality, Farrington and PPV adjusted |
| Non-Hemorrhagic stroke (risk interval days 22-42) | Seasonality, Farrington and PPV adjusted |
| Non-Hemorrhagic stroke or Transient ischemic attack (risk interval days 1-21) | Seasonality, Farrington and PPV adjusted |
| Non-Hemorrhagic stroke or Transient ischemic attack (risk interval days 22-42) | Seasonality, Farrington and PPV adjusted |
| Transient ischemic attack (risk interval days 1-21) | Seasonality and PPV adjusted |
| Transient ischemic attack (risk interval days 22-42) | Seasonality and PPV adjusted |
| Transverse myelitis | Seasonality adjusted |

**Supplementary Figure 1. Population eligibility requirements for descriptive and inferential analyses**

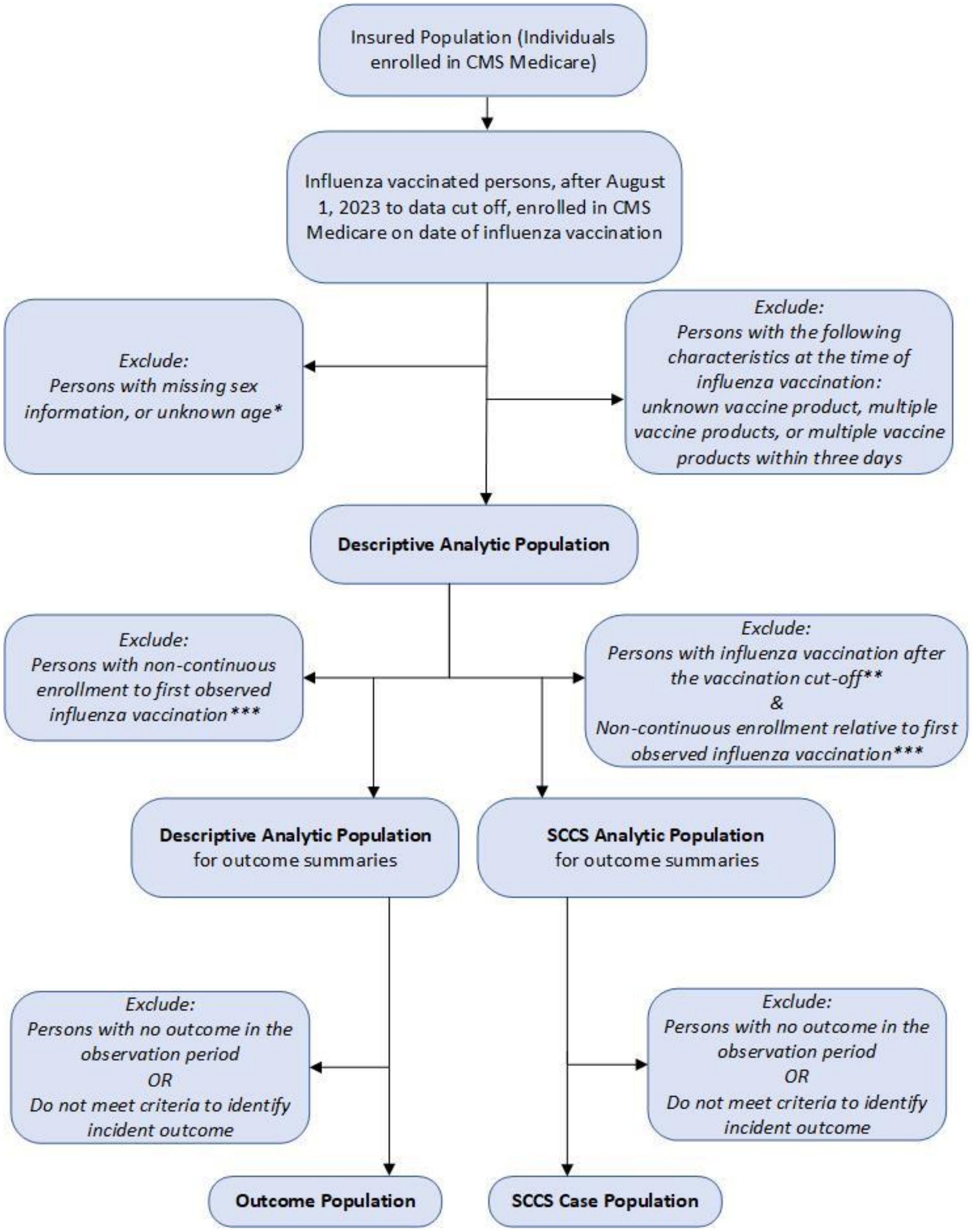


** Medicare beneficiaries must be aged 65 years and older on the date of their first vaccine dose for study inclusion.*

*** Cases eligible for the SCCS analysis will be determined by identifying the number of vaccine exposures whose expected observation period meet a 90% data-completeness threshold. This threshold is met if the last calendar day of the planned observation period is expected to have 90% or greater data-completeness based on the outcome-specific claims-delay distribution estimated from historical data.*

**** Medicare FFS beneficiaries will be required to have continuous enrollment in Medicare Parts A and B from 365 days prior to influenza vaccination. Medicare MA beneficiaries will be required to have continuous enrollment in Medicare Parts A, B, and C from 365 days prior to influenza vaccination.*

*Note: Medicare health plan beneficiaries will also require enrollment in a health plan with available prescription or pharmacy health plan data on vaccination date in the secondary analysis only, to ensure comprehensive capture of relevant concomitant immunizations.*
